# Long-term symptom profiles after COVID-19 *vs* other acute respiratory infections: a population-based observational study (COVIDENCE UK)

**DOI:** 10.1101/2023.07.06.23292296

**Authors:** Giulia Vivaldi, Paul E Pfeffer, Mohammad Talaei, Jayson Basera, Seif O Shaheen, Adrian R Martineau

**Author notes:** Correspondence to: Giulia Vivaldi, Blizard Institute, Barts and The London School of Medicine and Dentistry, Queen Mary University of London, London E1 2AT, UK.

## Abstract

**Background:** Long COVID is a well recognised, if heterogeneous, entity. Acute respiratory infections (ARIs) due to other pathogens may cause long-term symptoms, but few studies compare post-acute sequelae between SARS-CoV-2 and other ARIs. We aimed to compare symptom profiles between people with previous SARS-CoV-2 infection, people with previous non-COVID-19 ARIs, and contemporaneous controls, and to identify clusters of long-term symptoms.

**Methods:** COVIDENCE UK is a prospective, population-based UK study of ARIs in adults. We analysed data on 16 potential long COVID symptoms and health-related quality of life (HRQoL), reported in January, 2021, by participants unvaccinated against SARS-CoV-2. We classified participants as having previous SARS-CoV-2 infection or previous non-COVID-19 ARI (≥4 weeks prior) or no reported ARI. We compared symptoms by infection status using logistic and fractional regression, and identified symptom clusters using latent class analysis (LCA).

**Findings:** We included 10,203 participants (1343 [13.2%] with SARS-CoV-2 infection, 472 [4.6%] with non-COVID-19 ARI). Both types of infection were associated with increased prevalence/severity of most symptoms and decreased HRQoL compared with no infection. Participants with SARS-CoV-2 infection had increased odds of taste/smell problems and hair loss compared with participants with non-COVID-19 ARIs. Separate LCA models identified three symptom severity groups for each infection type. In the most severe groups (including 23% of participants with SARS-CoV-2, and 21% with non-COVID-19 ARI), SARS-CoV-2 infection presented with a higher probability of memory problems, difficulty concentrating, hair loss, and taste/smell problems than non-COVID-19 ARI.

**Interpretation:** Both SARS-CoV-2 and non-COVID-19 ARIs are associated with a wide range of long-term symptoms. Research on post-acute sequelae of ARIs should extend from SARS-CoV-2 to include other pathogens.

**Funding:** Barts Charity.

**Research in context:** *Evidence before this study:* We searched PubMed and Google Scholar for studies on post-acute sequelae of COVID-19 and other acute respiratory infections (ARIs), published up to May 24, 2023. We used search terms relating to COVID-19 and other ARIs (“COVID-19”, “SARS”, “severe acute respiratory syndrome”, “Middle East respiratory”, “MERS”, “respiratory infection”, “influenza”, “flu”) and post-acute symptoms (“long COVID”, “post-acute”, “PACS”, “sequelae”, “long-term”). Previous studies have shown a wide range of post-acute sequelae for COVID-19, affecting people with all severities of the acute disease. The few studies that have compared long-term symptoms between people with COVID-19 and non-COVID-19 ARIs have generally found a higher symptom burden among people with COVID-19; however, these studies have been restricted to hospitalised patients or electronic health record data, and thus do not capture the full picture in the community. Research into long COVID phenotypes has been inconclusive, with some analyses classifying people with long COVID according to the types of symptoms experienced, and others classifying them according to the overall severity of their symptoms.

*Added value of this study:* In this population-based study of ARIs in the community, we observed high symptom burden among people with previous SARS-CoV-2 infection when compared with controls, highlighting the extensive reach of long COVID. Our finding of a similar symptom burden among people with non-COVID-19 ARIs suggests that post-acute sequelae of other ARIs may be going unrecognised, particularly given that the vast majority did not experience a severe acute infection. Latent class analyses of symptoms identified groupings based on overall symptom severity, rather than symptom types, for both SARS-CoV-2 infections and non-COVID-19 ARIs, suggesting that overall symptom burden may best characterise the experience of people with post-acute sequelae. Notably, among participants with the most severe symptoms, only half of those with previous SARS-CoV-2 infection attributed their symptoms to long COVID, suggesting they either did not believe the infection was the cause, or they did not consider their symptoms severe enough to qualify as long COVID.

*Implications of all the available evidence:* The long-term symptoms experienced by some people with previous ARIs, including SARS-CoV-2, highlights the need for improved understanding, diagnosis, and treatment of post-acute infection syndromes. As much-needed research into long COVID continues, we must take the opportunity to investigate and consider the post-acute burden of ARIs due to other pathogens.

## Introduction

Long COVID—the post-acute sequelae of SARS-CoV-2 infection—is a highly heterogeneous condition. It can be broadly defined as new or ongoing symptoms more than 4 weeks after the acute infection,^1^ with some of the most common being fatigue, dyspnoea, and cognitive impairment.^2–5^ However, more than 200 symptoms have been investigated,^4^ making it difficult to precisely characterise, diagnose, or treat the condition.^6^ Its high heterogeneity and multisystem nature has encouraged the search for different long COVID phenotypes, with the aim of improving understanding of the condition and targeting treatment options for different phenotypes.

Long COVID is estimated to affect at least 10% of people infected with SARS-CoV-2, with far higher incidence among those hospitalised.^6^ With more than 670 million people infected to date,^7^ the number of people globally affected by long COVID may be estimated at 67 million, and will almost certainly be higher.^6^ Importantly, this high incidence of long COVID has taken place over a short period of time—the 3 years of the COVID-19 pandemic—meaning that awareness of these post-acute sequelae has grown much more rapidly than for other coronavirus pandemics with smaller reach and a lower percentage of survivors.^8, 9^

Post-acute infection syndromes are not a new phenomenon; indeed, many cases of chronic fatigue syndrome are reported to follow an infection-like episode.^10^ Nonetheless, these syndromes often go undiagnosed owing to the wide range of symptoms and lack of diagnostic tests.^10^ Studies into Middle East respiratory syndrome, SARS-CoV-1, and influenza have identified chronic pulmonary and extrapulmonary sequelae a year after the acute infection,^8^ but research has largely been restricted to people with severe disease. By contrast, long COVID has been studied in people with all levels of severity of the acute infection,^2, 6, 11^ and has been found to affect even those with mild initial symptoms.^11^

This spotlight on the long-term effects of SARS-CoV-2 infection leads to the question of whether there are post-acute sequelae of other acute respiratory infections (ARIs) in the community, including those that present with mild initial symptoms, and how these sequelae compare with long COVID. Importantly, given the non-specific nature of symptoms such as fatigue, any comparison needs to take into account the symptom prevalence in a contemporaneous, uninfected population— particularly given the background of a disruptive pandemic, with increased stressors, limitations on movements, and elevated levels of population fatigue.^12^ We therefore aimed to compare symptom profiles between people with previous SARS-CoV-2 infection, people with previous non-COVID-19 ARIs, and those with no reported infection, and then to identify distinct symptom clusters among people with previous SARS-CoV-2 infections or non-COVID-19 ARIs.

## Methods

### Study participants

COVIDENCE UK is a prospective, longitudinal, population-based observational study of COVID-19 in the UK population (https://www.qmul.ac.uk/covidence). Inclusion criteria were age 16 years or older and UK residence at enrolment, with no exclusion criteria. Participants were invited via a national media campaign to complete an online baseline questionnaire and monthly follow-up questionnaires to capture information on potential symptoms of COVID-19, results of nose or throat swab tests for SARS-CoV-2, SARS-CoV-2 vaccine status, and details of a wide range of potential long COVID symptoms. The study was launched on May 1, 2020, and closed to enrolment on Oct 6, 2021. This study is registered with ClinicalTrials.gov, NCT04330599.

For this analysis, we included all COVIDENCE UK participants responding to the January, 2021, follow-up questionnaire who had not been vaccinated against SARS-CoV-2. We focused on this date for several reasons. During this period, all COVIDENCE UK participants were asked a panel of questions about potential long COVID symptoms, which allowed us to assess prevalence of potential long COVID symptoms regardless of reported previous infections or self-reported long COVID. Additionally, January, 2021, was sufficiently early in the vaccine rollout that most of our participants were not vaccinated, and sufficiently late into the pandemic that we were able to record a substantial number of infections.

Participants were categorised as having had a previous SARS-CoV-2 infection, a previous non-COVID-19 ARI, or no recorded infections during follow-up. Participants who reported both a previous SARS-CoV-2 infection and a previous non-COVID-19 ARI were excluded. To minimise infection misclassification, we also excluded participants who reported symptom-defined non-COVID-19 ARIs that were not accompanied by a negative SARS-CoV-2 swab test (lateral flow or RT-PCR). Finally, we excluded any participants who had either infection fewer than 4 weeks before the survey date, in order to focus on long-term symptoms.

### Previous infections

We defined previous SARS-CoV-2 infection as any of the following: a positive swab test for SARS-CoV-2, a positive antibody test for SARS-CoV-2, or symptom-defined probable COVID-19, based on the algorithm described by Menni et al.^13^ Infection dates were defined as the date of the test, for swab tests, and the date of symptom onset, for participants with symptom-defined COVID-19. Participants reporting positive antibody tests were asked to recall a suspected infection date (based on symptoms, swab test results, or close proximity to a COVID-19 case); participants unable to provide a date who nonetheless reported probable COVID-19 symptoms^13^ before their antibody test were assigned the symptom onset date as their infection date. Participants with positive swab or antibody tests and no reported symptoms were defined as asymptomatic cases.

We defined a previous non-COVID-19 ARI as self-report of a general practitioner or hospital diagnosis of pneumonia, influenza, bronchitis, tonsillitis, pharyngitis, ear infection, common cold, or other upper or lower respiratory infection not caused by SARS-CoV-2; or self-report of a symptom-defined ARI accompanied by a negative SARS-CoV-2 swab test (lateral flow or RT-PCR). Symptom-defined episodes of ARI were identified using modified Jackson criteria for upper respiratory infection,^14^ modified Macfarlane criteria for lower respiratory infection,^15^ and a triad of cough, fever, and myalgia for influenza-like illness.^16^ Infection dates were defined as the date of symptom onset.

### Symptoms

We assessed the prevalence or severity of 16 potential COVID-19 symptoms: coughing, problems with sleep, memory problems, difficulty concentrating, muscle or joint pain, problems with sense of taste or smell, diarrhoea, stomach problems (abdominal pains), changes to voice, hair loss, unusual racing of the heart, lightheadedness or dizziness, unusual sweating, breathlessness, anxiety or depression, and fatigue.^17^ We measured breathlessness with the Medical Research Council (MRC) Dyspnoea Scale,^18^ anxiety or depression with the Patient Health Questionnaire-4 (PHQ-4),^19^ and fatigue with the Functional Assessment of Chronic Illness Therapy (FACIT) Fatigue Score.^20^ For symptoms measured on a scale (dyspnoea, anxiety or depression, and fatigue), we refer to the severity of symptoms; for all other symptoms, we refer to the prevalence (ie, whether they were present or absent).

We also assessed health-related quality of life (HRQoL) using the EQ-5D-3L,^21^ which covers five dimensions of health—mobility, self-care, usual activities, pain or discomfort, and anxiety or depression—and includes a visual analogue scale (VAS) to record the respondent’s self-rated health status on a scale from 0 to 100.

### Statistical analysis

All analyses considered symptoms at a single timepoint. For our first analysis, we compared prevalence or severity of potential long COVID symptoms and HRQoL between participants with previous SARS-CoV-2 infection, those with non-COVID-19 ARIs, and those with no reported infections. To explore the long-term effects of COVID-19, we additionally compared symptoms between participants with a SARS-CoV-2 infection more than 12 weeks prior and those with no infection. When comparing previous SARS-CoV-2 infection with non-COVID-19 ARIs, we restricted the analysis to participants with symptomatic SARS-CoV-2 infection, as participants had no means of reporting asymptomatic non-COVID-19 ARIs. Finally, we explored how symptoms differed among participants with previous SARS-CoV-2 infection according to time since infection (4–12 weeks *vs* >12 weeks) and self-reported severity of the initial infection, classed as either asymptomatic (no symptoms reported), mild (able to do most of usual activities), moderate (unable to do usual activities, but without requiring bedrest), severe (requiring bedrest), and hospitalised.

Regressions were carried out in all participants, adjusted for potential confounders that could influence both risk or severity of infection and likelihood of experiencing or reporting symptoms: age, sex, highest educational attainment, baseline self-reported general health, body-mass index, asthma or chronic obstructive pulmonary disease, and having received an influenza vaccination in the previous 6 months. Categorical variables were analysed using logistic regression or ordered logistic regression, as appropriate. The proportional odds assumption for ordinal variables (MRC Dyspnoea, PHQ-4, EQ-5D Activities, and EQ-5D Pain) was assessed using the Brant test.^22^ Models with significant deviations were refitted using a partial proportional odds model;^23^ as deviations are common with large sample sizes, and proportional odds models are less parsimonious, model fit was compared using Lacy’s adjusted ordinal explained variation measure.^24^ Continuous, bounded variables (FACIT-13 and the EQ-5D Visual Analogue Scale) were rescaled to the [0,1] interval and analysed using fractional regression. Regression results are presented as odds ratios for both types of logistic regression and as predicted percentage point change in outcome for fractional regressions, to aid interpretation. Further details on these analyses can be found in the appendix (pp 3–4). We adjusted for multiple testing using the Benjamini–Hochberg procedure.^25^

We carried out three sensitivity analyses: first, we repeated the analyses in participants who had swab-test or antibody-test confirmed COVID-19 (ie, excluding any participants with symptom-defined potential COVID-19). Second, to better account for baseline health differences that may have influenced susceptibility to infection, we repeated analyses by infection status excluding those whose reported infections occurred before cohort enrolment, so that the cohort baseline general health would represent health status before infection. Third, to compare the long-term effects of SARS-CoV-2 and non-COVID-19 ARIs, we repeated the comparison between participants with previous infections restricted to participants whose infections occurred more than 12 weeks prior.

For our second analysis, we carried out separate latent class analyses (LCA) in participants who had previous SARS-CoV-2 infection or a previous non-COVID-19 ARI, to explore whether there was any clustering of symptoms. LCA is a statistical method of identifying subgroups in a given population who share measured characteristics, such as certain symptoms. The underlying assumption is that different patterns of symptoms can be explained by membership to a certain group or class. With this approach, we hoped to identify distinct symptom phenotypes among people who had a previous SARS-CoV-2 infection or a non-COVID-19 ARI.

We fitted latent class models with up to ten classes, and identified the optimal number of classes by considering changes in the Bayesian Information Criterion (BIC), the Vuong-Lo-Mendell-Rubin (VLMR) adjusted likelihood ratio test, and the bootstrapped likelihood ratio test.^26^ We additionally considered the interpretability and separation of any identified classes, as well as examining classification diagnostics such as model entropy and average latent class posterior probabilities.^26^ Once the number of classes had been chosen, we attempted to improve model fit by considering direct effects between pairs of indicators that showed violations in the independence assumption. Finally, we adjusted for age and sex in the final model by including them as covariates.^27^ Owing to problems with convergence, we converted responses to the five dimensions of the EQ-5D-3L into a weighted health state index, using EQ-5D preference weights obtained from the UK general population.^28^ The upper bound of 1 represents full health, whereas a value of 0 represents death. Negative values are allowed (ie, states considered to be worse than death).

Analyses were done in Stata (version 17.0) and LatentGOLD (version 6.0).

## Results

10,203 participants were included in the analysis (appendix figure S1), with a median age of 62.8 years (IQR 53.7–68.9). 6994 (68.6%) participants were female and the vast majority were White (table 1). 1343 (13.2%) had a previous SARS-CoV-2 infection and 472 (4.6%) had a previous non-COVID-19 ARI (table 1). 241 (18.1%) of participants with previous SARS-CoV-2 infection reported a positive swab test and 324 (24.4%) a positive antibody test; the remaining participants had symptom-defined infections. The majority of participants with previous SARS-CoV-2 infection were infected more than 12 weeks earlier, and 40 (6.7%) were hospitalised (table 1). Participants with no infection were the least likely to smoke or vape, whereas participants with non-COVID-19 ARIs were the most likely to have asthma (table 1). Symptom prevalence by infection status, timing, and severity is shown in the appendix (figures S2–S4).

**Table 1:** Participant characteristics.

|  | Previous SARS-CoV-2 infection (N=1,343) | Previous non-COVID-19 ARI (N=472) | No infection (N=8,388) |
| --- | --- | --- | --- |
| **Sociodemographics** |  |  |  |
| Age, years | 59.4 (50.8–66.2) | 57.2 (47.4–65.0) | 63.5 (54.8–69.3) |
| <30 | 43 (3.2%) | 30 (6.4%) | 242 (2.9%) |
| 30 to <40 | 86 (6.4%) | 44 (9.3%) | 411 (4.9%) |
| 40 to <50 | 181 (13.5%) | 74 (15.7%) | 792 (9.4%) |
| 50 to <60 | 392 (29.2%) | 137 (29.0%) | 1,716 (20.5%) |
| 60 to <70 | 467 (34.8%) | 136 (28.8%) | 3,361 (40.1%) |
| ≥70 | 174 (13.0%) | 51 (10.8%) | 1,866 (22.2%) |
| Sex |  |  |  |
| Female | 924 (68.8%) | 385 (81.6%) | 5,685 (67.8%) |
| Male | 419 (31.2%) | 87 (18.4%) | 2,703 (32.2%) |
| Ethnicity |  |  |  |
| White | 1,259 (93.7%) | 453 (96.0%) | 8,010 (95.5%) |
| Mixed/multiple/other ethnic groups | 46 (3.4%) | 10 (2.1%) | 206 (2.5%) |
| South Asian | 27 (2.0%) | 7 (1.5%) | 121 (1.4%) |
| Black/African/Caribbean/Black British | 11 (0.8%) | 2 (0.4%) | 51 (0.6%) |
| Country |  |  |  |
| England | 1,207 (89.9%) | 430 (91.1%) | 7,400 (88.3%) |
| Northern Ireland | 20 (1.5%) | 11 (2.3%) | 144 (1.7%) |
| Scotland | 73 (5.4%) | 23 (4.9%) | 533 (6.4%) |
| Wales | 43 (3.2%) | 8 (1.7%) | 306 (3.7%) |
| No. people per bedroom |  |  |  |
| <1 | 826 (61.9%) | 278 (59.1%) | 5,849 (70.1%) |
| 1 to <2 | 472 (35.4%) | 173 (36.8%) | 2,341 (28.1%) |
| 2 to <3 | 33 (2.5%) | 17 (3.6%) | 145 (1.7%) |
| 3 to <4 | 3 (0.2%) | 2 (0.4%) | 4 (0.0%) |
| Quartiles of IMD decile |  |  |  |
| Q4 (least deprived) | 431 (32.1%) | 145 (30.7%) | 2,737 (32.7%) |
| Q3 | 299 (22.3%) | 103 (21.8%) | 2,259 (27.0%) |
| Q2 | 276 (20.6%) | 122 (25.8%) | 1,699 (20.3%) |
| Q1 (most deprived) | 336 (25.0%) | 102 (21.6%) | 1,683 (20.1%) |
| Frontline worker |  |  |  |
| No | 1,091 (81.2%) | 374 (79.2%) | 7,367 (87.9%) |
| Non-health | 190 (14.1%) | 84 (17.8%) | 838 (10.0%) |
| Health | 62 (4.6%) | 14 (3.0%) | 173 (2.1%) |
| Highest educational level attained |  |  |  |
| Primary or secondary | 158 (11.8%) | 54 (11.5%) | 950 (11.3%) |
| Higher or further (A levels) | 187 (13.9%) | 71 (15.1%) | 1,279 (15.3%) |
| College or university | 605 (45.0%) | 190 (40.3%) | 3,682 (43.9%) |
| Post-graduate | 393 (29.3%) | 156 (33.1%) | 2,467 (29.4%) |
| **Clinical characteristics** |  |  |  |
| BMI, kg/m<sup>2</sup> | 25.5 (22.9–29.2) | 25.5 (23.3–29.3) | 24.9 (22.5–28.3) |
| <25 | 603 (44.9%) | 213 (45.3%) | 4,256 (50.8%) |
| 25 to <30 | 455 (33.9%) | 155 (33.0%) | 2,632 (31.4%) |
| ≥30 | 284 (21.2%) | 102 (21.7%) | 1,487 (17.8%) |
| Asthma | 269 (20.0%) | 114 (24.2%) | 1,239 (14.8%) |
| Atopy | 359 (26.7%) | 136 (28.8%) | 1,958 (23.3%) |
| Autoimmune disease | 113 (8.4%) | 48 (10.2%) | 708 (8.4%) |
| Cancer |  |  |  |
| Past (cured or in remission) | 107 (8.0%) | 34 (7.2%) | 739 (8.8%) |
| Active treatment | 10 (0.7%) | 5 (1.1%) | 73 (0.9%) |
| COPD | 38 (2.8%) | 8 (1.7%) | 156 (1.9%) |
| Diabetes | 42 (3.1%) | 21 (4.4%) | 393 (4.7%) |
| Heart disease | 36 (2.7%) | 22 (4.7%) | 293 (3.5%) |
| Hypertension | 260 (19.4%) | 73 (15.5%) | 1,803 (21.5%) |
| Immunodeficiency | 13 (1.0%) | 2 (0.4%) | 43 (0.5%) |
| Kidney disease | 26 (1.9%) | 12 (2.5%) | 142 (1.7%) |
| Major neurological conditions | 40 (3.0%) | 9 (1.9%) | 202 (2.4%) |
| **Infections** |  |  |  |
| Weeks since infection |  |  |  |
| 4–12 weeks | 166 (12.4%) | 274 (58.1%) | .. |
| >12 weeks | 1,177 (87.6%) | 198 (41.9%) | .. |
| Infection severity* |  |  |  |
| Asymptomatic | 90/757 (11.9%) | .. | .. |
| Mildly unwell | 161/757 (21.3%) | 317 (67.2%) | .. |
| Moderately unwell | 178/757 (23.5%) | 89 (18.9%) | .. |
| Very unwell | 288/757 (38.0%) | 58 (12.3%) | .. |
| Hospitalised | 40/757 (5.3%) | 8 (1.7%) | .. |

ARI=acute respiratory infection. BMI=body-mass index. COPD=chronic obstructive pulmonary disease. IMD=Index of Multiple Deprivation. \*For symptomatic and non-hospitalised participants, severity was self-reported with the following statements: “Mildly unwell – I could do most of my usual activities”, “Moderately unwell – I couldn’t do usual activities but didn’t need to go to bed in the daytime”, and “Very unwell – I had to go to bed in the daytime”.

We observed increased prevalence or severity of all symptoms, accompanied by decreased HRQoL, among participants with previous SARS-CoV-2 infection compared with those without any infection, regardless of whether their infection had been more than 12 weeks prior (table 2). The greatest difference observed was in the prevalence of problems with taste or smell, although this was slightly reduced among participants who had been infected more than 12 weeks prior (table 2). When compared with participants with no infection, those with non-COVID-19 ARIs also showed increased prevalence or severity of most symptoms considered, with the exception of muscle or joint pain, problems with sense of taste or smell, and hair loss (table 2).

**Table 2:** Symptom associations among participants with previous SARS-CoV-2 infection *vs* no infection or non-COVID-19 ARI.

|  | Previous SARS-CoV-2 infection vs no infection |  | SARS-CoV-2 infection >12 weeks prior vs no infection |  | Previous SARS-CoV-2 infection vs previous non-COVID-19 ARI |  | Previous non-COVID-19 ARI vs no infection |  |
| --- | --- | --- | --- | --- | --- | --- | --- | --- |
|  | Estimate* (95% CI) | p value† | Estimate* (95% CI) | p value† | Estimate* (95% CI) | p value‡ | Estimate* (95% CI) | p value§ |
| Coughing | 2.01 (1.69–2.38) | <0.001 | 1.78 (1.48–2.15) | <0.001 | 0.75 (0.57–0.99) | 0.041 | 2.90 (2.27–3.71) | <0.001 |
| Problems with sleep | 1.27 (1.13–1.44) | <0.001 | 1.27 (1.12–1.45) | <0.001 | 0.82 (0.66–1.03) | 0.089 | 1.51 (1.25–1.84) | <0.001 |
| Memory problems | 2.08 (1.80–2.41) | <0.001 | 2.06 (1.76–2.40) | <0.001 | 1.34 (1.03–1.75) | 0.032 | 1.65 (1.29–2.09) | <0.001 |
| Difficulty concentrating | 1.92 (1.66–2.23) | <0.001 | 1.90 (1.63–2.22) | <0.001 | 1.24 (0.96–1.60) | 0.100 | 1.56 (1.24–1.97) | <0.001 |
| Muscle or joint pain | 1.35 (1.19–1.54) | <0.001 | 1.34 (1.17–1.54) | <0.001 | 1.13 (0.89–1.43) | 0.311 | 1.22 (1.00–1.50) | 0.055 |
| Problems with sense of smell / taste | 9.25 (7.45–11.48) | <0.001 | 8.23 (6.55–10.34) | <0.001 | 8.06 (4.52–14.35) | <0.001 | 1.28 (0.71–2.29) | 0.414 |
| Diarrhoea | 1.74 (1.42–2.13) | <0.001 | 1.60 (1.28–1.99) | <0.001 | 0.86 (0.62–1.19) | 0.357 | 2.11 (1.58–2.82) | <0.001 |
| Stomach (abdominal pains) | 1.73 (1.43–2.08) | <0.001 | 1.73 (1.42–2.10) | <0.001 | 0.85 (0.63–1.14) | 0.278 | 2.13 (1.64–2.78) | <0.001 |
| Changes to voice | 2.70 (2.07–3.51) | <0.001 | 2.49 (1.89–3.30) | <0.001 | 0.84 (0.56–1.26) | 0.402 | 3.32 (2.29–4.80) | <0.001 |
| Hair loss | 2.07 (1.69–2.54) | <0.001 | 2.08 (1.68–2.57) | <0.001 | 2.10 (1.39–3.19) | <0.001 | 1.05 (0.71–1.53) | 0.821 |
| Unusual racing of the heart | 2.24 (1.86–2.70) | <0.001 | 2.17 (1.78–2.64) | <0.001 | 1.25 (0.91–1.73) | 0.175 | 1.81 (1.35–2.42) | <0.001 |
| Lightheadedness or dizziness | 2.16 (1.84–2.54) | <0.001 | 2.10 (1.77–2.49) | <0.001 | 1.34 (1.00–1.80) | 0.049 | 1.59 (1.22–2.07) | <0.001 |
| Unusual sweating | 2.44 (2.01–2.97) | <0.001 | 2.41 (1.96–2.96) | <0.001 | 1.31 (0.93–1.85) | 0.123 | 1.91 (1.40–2.63) | <0.001 |
| MRC Dyspnoea \|\| | 1.30 (1.14–1.49) | <0.001 | 1.20 (1.04–1.38) | 0.014 | 0.97 (0.77–1.22) | 0.780 | 1.39 (1.13–1.72) | 0.002 |
| PHQ-4 grade \|\| | 1.30 (1.13–1.49) | <0.001 | 1.25 (1.08–1.45) | 0.003 | 0.92 (0.73–1.17) | 0.489 | 1.34 (1.09–1.65) | 0.006 |
| FACIT-13 score (reversed)¶ | 3.5% (2.6–4.3) | <0.001 | 3.0% (2.1–3.9) | <0.001 | -0.3% (-2.1–1.5) | 0.730 | 3.6% (2.3–4.9) | <0.001 |
| EQ-5D Activities \|\| | 2.05 (1.71–2.46) | <0.001 | 1.95 (1.61–2.36) | <0.001 | 1.03 (0.77–1.39) | 0.821 | 1.92 (1.46–2.54) | <0.001 |
| EQ-5D Pain \|\| | 1.39 (1.21–1.58) | <0.001 | 1.39 (1.21–1.60) | <0.001 | 0.98 (0.77–1.24) | 0.874 | 1.41 (1.14–1.74) | 0.002 |
| EQ-5D Self-care (two groups) | 1.14 (0.81–1.59) | 0.460 | 1.22 (0.87–1.73) | 0.253 | 1.29 (0.72–2.33) | 0.391 | 0.98 (0.58–1.65) | 0.940 |
| EQ-5D Mobility (two groups) | 1.08 (0.88–1.32) | 0.454 | 1.03 (0.83–1.28) | 0.790 | 0.89 (0.64–1.25) | 0.515 | 1.24 (0.91–1.69) | 0.173 |
| EQ-5D VAS¶ | -1.9% (-2.9 to -1.0) | <0.001 | -1.7% (-2.6 to -0.7) | 0.001 | 0.1% (-1.7 to 2.0) | 0.888 | -1.6% (-3.1 to 0.0) | 0.043 |

Comparison between non-COVID-19 ARI and no infection is shown in the main paper. ARI=acute respiratory infection. PHQ=Patient Health Questionnaire. MRC=Medical Research Council. VAS=visual analogue scale. \*Estimates are odds ratios for binary and ordinal outcomes, and predicted percentage point changes for continuous outcomes. †After adjustment for multiple corrections, p<0.04 is the threshold for statistical significance. ‡After adjustment for multiple corrections, p<0.005 is the threshold for statistical significance. §After adjustment for multiple testing, p<0.01 is the threshold for statistical significance. || Ordinal outcome. ¶Continuous outcome. Raw coefficients are available in the appendix (table S1).

When compared with participants with non-COVID-19 ARI, those with SARS-CoV-2 infection had greater odds of reporting problems with sense of taste or smell and hair loss, but few other differences were observed (table 2).

We found that participants with SARS-CoV-2 infection who had been infected more than 12 weeks prior were less likely to report coughing or problems with taste or smell, and had lower levels of dyspnoea and fatigue, than those infected 4–12 weeks prior (table 3).

**Table 3:** Symptom associations by time since infection, among participants with previous SARS-CoV-2 infection.

|  | SARS-CoV-2 infection >12 weeks prior vs<br>4–12 weeks prior |  |
| --- | --- | --- |
|  | Estimate* (95% CI) | p value† |
| Coughing | 0.43 (0.29–0.65) | <0.001 |
| Problems with sleep | 1.01 (0.72–1.42) | 0.958 |
| Memory problems | 0.91 (0.61–1.36) | 0.637 |
| Difficulty concentrating | 0.90 (0.61–1.33) | 0.594 |
| Muscle or joint pain | 0.98 (0.68–1.40) | 0.890 |
| Problems with sense of smell / taste | 0.42 (0.28–0.63) | <0.001 |
| Diarrhoea | 0.58 (0.36–0.95) | 0.031 |
| Stomach (abdominal pains) | 1.07 (0.64–1.80) | 0.802 |
| Changes to voice | 0.50 (0.27–0.94) | 0.030 |
| Hair loss | 1.01 (0.57–1.78) | 0.986 |
| Unusual racing of the heart | 0.77 (0.48–1.24) | 0.276 |
| Lightheadedness or dizziness | 0.82 (0.54–1.24) | 0.341 |
| Unusual sweating | 0.84 (0.50–1.39) | 0.493 |
| MRC Dyspnoea‡ | 0.57 (0.40–0.81) | 0.002 |
| PHQ-4 grade‡ | 0.79 (0.55–1.14) | 0.206 |
| FACIT-13 score (reversed)§ | -4.1% (-7.3 to -1.0) | 0.009 |
| EQ-5D Activities‡ | 0.74 (0.47–1.17) | 0.197 |
| EQ-5D Pain‡ | 1.06 (0.73–1.53) | 0.777 |
| EQ-5D Self-care (two groups) | 3.10 (0.71–13.55) | 0.132 |
| EQ-5D Mobility (two groups) | 0.74 (0.44–1.26) | 0.264 |
| EQ-5D VAS§ | 2.2% (-0.7 to 5.1) | 0.137 |

PHQ=Patient Health Questionnaire. MRC=Medical Research Council. VAS=visual analogue scale. \*Estimates are odds ratios for binary and ordinal outcomes, and predicted percentage point changes for continuous outcomes. †After adjustment for multiple corrections, $p<0.01$ is the threshold for statistical significance. ‡Ordinal outcome. §Continuous outcome. Raw coefficients are available in the appendix (table S1).

We observed a link between the severity of the initial infection and subsequent symptom prevalence (table 4). Compared with participants with an asymptomatic SARS-CoV-2 infection, those who reported mild severity of their initial infection had increased odds of problems with sense of taste or smell, whereas those who were moderately unwell had increased odds of coughing (table 4). Participants who were very unwell had increased odds of memory problems, difficulty concentrating, and problems with sense of taste or smell, as well as higher levels of fatigue (table 4). Participants who had been hospitalised had increased prevalence or severity of nine symptoms or HRQoL measures (table 4). In particular, previously hospitalised participants had substantially increased odds of reporting difficulty concentrating, memory problems, and problems with everyday activities (table 4). Compared with asymptomatic participants, their reported levels of fatigue were 21 percentage points higher (table 4).

**Table 4:** Symptom associations by severity of infection, among participants with previous SARS-CoV-2 infection.

|  | Mildly unwell vs asymptomatic |  | Moderately unwell vs asymptomatic |  | Very unwell vs asymptomatic |  | Hospitalised vs asymptomatic |  |
| --- | --- | --- | --- | --- | --- | --- | --- | --- |
|  | Estimate* (95% CI) | p value† | Estimate* (95% CI) | p value‡ | Estimate* (95% CI) | p value§ | Estimate* (95% CI) | p value\|\| |
| Coughing | 2.16 (0.77–6.07) | 0.144 | 4.80 (1.80–12.76) | 0.002 | 4.05 (1.55–10.56) | 0.004 | 3.37 (1.00–11.39) | 0.051 |
| Problems with sleep | 0.73 (0.42–1.27) | 0.263 | 1.04 (0.61–1.78) | 0.894 | 0.98 (0.59–1.61) | 0.924 | 1.93 (0.83–4.48) | 0.126 |
| Memory problems | 1.57 (0.71–3.50) | 0.265 | 2.34 (1.09–5.01) | 0.029 | 3.63 (1.76–7.49) | <0.001 | 11.97 (4.49–31.91) | <0.001 |
| Difficulty concentrating | 2.44 (1.07–5.57) | 0.034 | 3.15 (1.42–7.02) | 0.005 | 4.37 (2.04–9.37) | <0.001 | 16.23 (5.82–45.24) | <0.001 |
| Muscle or joint pain | 0.96 (0.55–1.71) | 0.902 | 1.08 (0.62–1.90) | 0.778 | 1.60 (0.95–2.69) | 0.077 | 3.16 (1.34–7.42) | 0.008 |
| Problems with sense of smell / taste | 5.19 (1.94–13.83) | 0.001 | 4.14 (1.55–11.04) | 0.004 | 4.87 (1.88–12.59) | 0.001 | 6.78 (2.14–21.48) | 0.001 |
| Diarrhoea | 2.05 (0.64–6.51) | 0.225 | 2.76 (0.90–8.39) | 0.074 | 3.03 (1.04–8.83) | 0.043 | 7.87 (2.24–27.69) | 0.001 |
| Stomach (abdominal pains) | 1.74 (0.65–4.66) | 0.274 | 2.09 (0.81–5.41) | 0.127 | 2.23 (0.90–5.51) | 0.083 | 7.30 (2.42–21.99) | <0.001 |
| Changes to voice | 2.14 (0.57–8.00) | 0.260 | 1.46 (0.38–5.57) | 0.577 | 2.21 (0.63–7.74) | 0.215 | 4.07 (0.89–18.56) | 0.070 |
| Hair loss | 0.75 (0.28–1.99) | 0.560 | 1.05 (0.42–2.59) | 0.919 | 1.40 (0.61–3.19) | 0.429 | 1.91 (0.61–6.00) | 0.265 |
| Unusual racing of the heart | 0.61 (0.25–1.50) | 0.285 | 0.80 (0.35–1.82) | 0.598 | 1.18 (0.57–2.48) | 0.654 | 4.24 (1.61–11.19) | 0.003 |
| Lightheadedness or dizziness | 0.78 (0.36–1.70) | 0.537 | 1.95 (0.97–3.90) | 0.059 | 1.70 (0.88–3.31) | 0.115 | 3.57 (1.44–8.84) | 0.006 |
| Unusual sweating | 1.06 (0.42–2.68) | 0.899 | 1.14 (0.47–2.76) | 0.776 | 2.17 (0.97–4.86) | 0.060 | 1.53 (0.48–4.86) | 0.474 |
| MRC Dyspnoea¶ | 1.48 (0.80–2.74) | 0.217 | 1.33 (0.72–2.45) | 0.357 | 1.73 (0.98–3.06) | 0.058 | 5.99 (2.68–13.37) | <0.001 |
| PHQ-4 grade¶ | 1.01 (0.54–1.89) | 0.973 | 1.18 (0.64–2.17) | 0.590 | 1.40 (0.79–2.46) | 0.247 | 2.68 (1.17–6.12) | 0.020 |
| FACIT-13 score (reversed)** | 1.2% (-2.6 to 5.0) | 0.533 | 1.4% (-2.3 to 5.1) | 0.462 | 6.2% (2.3–21.0) | 0.002 | 21.0% (12.3–29.6) | <0.001 |
| EQ-5D Activities¶ | 1.33 (0.56–3.14) | 0.514 | 1.87 (0.83–4.22) | 0.132 | 2.32 (1.07–5.01) | 0.032 | 12.58 (4.60–34.41) | <0.001 |
| EQ-5D Pain¶ | 0.84 (0.47–1.49) | 0.551 | 0.87 (0.50–1.53) | 0.636 | 1.01 (0.60–1.70) | 0.974 | 2.22 (0.98–5.00) | 0.055 |
| EQ-5D Mobility (two groups) | 1.91 (0.71–5.08) | 0.198 | 1.29 (0.48–3.46) | 0.609 | 1.77 (0.70–4.47) | 0.224 | 7.90 (2.55–24.49) | <0.001 |
| EQ-5D VAS** | -3.1% (-7.7 to 1.5) | 0.182 | 0.5% (-3.9 to 4.8) | 0.837 | -2.4% (-6.6 to 1.8) | 0.260 | -8.0% (-14.8 to -1.1) | 0.022 |

Differences in EQ-5D Self Care were not estimable due to small numbers. QPHQ=Patient Health Questionnaire. MRC=Medical Research Council. VAS=visual analogue scale. \*Estimates are odds ratios for binary and ordinal outcomes, and predicted percentage point changes for continuous outcomes. †After adjustment for multiple corrections, $p<0.002$ is the threshold for statistical significance. ‡After adjustment for multiple corrections, $p<0.005$ is the threshold for statistical significance. §After adjustment for multiple corrections, $p<0.01$ is the threshold for statistical significance. || After adjustment for multiple corrections, $p<0.04$ is the threshold for statistical significance. ¶Ordinal outcome. \*\*Continuous outcome. Raw coefficients are available in the appendix (table S1).

We obtained similar results in sensitivity analyses excluding participants with symptom-defined SARS-CoV-2 infection, although some analyses were affected by a lack of power (appendix tables S2– S4). When adjusting for pre-infection general health, fewer differences were observed between participants with SARS-CoV-2 infection more than 12 weeks prior and those with no infection, but odds of reporting problems with sense of taste or smell increased substantially, from 8.23 (95% CI 6.55–10.34) to 13.94 (8.47–22.93; table S6). Restricting analyses to participants with infections more than 12 weeks prior did not change our results, although the analysis was affected by a lack of power (appendix table S7). Finally, we did an exploratory analysis to compare symptom prevalence and severity between participants with asymptomatic or mild SARS-CoV-2 infection and those with no infection, to explore how much our results were driven by participants with a more severe acute infection; while the odds ratios were lower, we continued to find significant differences between the two groups for all symptoms and measures, except for dyspnoea, self-care, and mobility (data not shown).

For the LCA in participants with previous SARS-CoV-2 infection, our final model was a three-class model, with an entropy of 0.948 and a strong distinction between classes (appendix table S9); further details on model selection are given in the appendix (pp 14–16). The three classes can be interpreted as different levels of ongoing symptom severity: mild (comprising 45% of participants), moderate (32% of participants), and severe (23% of participants). Figure 1A shows the symptom profile for these three classes. The mild class presents with generally low prevalence and severity of the symptoms considered, with the most frequent symptoms being sleep problems and muscle or joint pain (figure 1). Participants assigned to the moderate class present with a slightly increased probability of reporting all symptoms, but with a larger increase in muscle or joint pain, sleep problems, memory problems, difficulty concentrating, and PHQ-4 score (figure 1A). The final class, comprising nearly a quarter of participants with previous SARS-CoV-2 infection, shows a marked increase in the prevalence and severity of all symptoms, with the greatest increases among memory problems, difficulty concentrating, and lightheadedness or dizziness (figure 1A). When comparing participant characteristics across the three classes, we observed that with increasing symptom severity, participants were more likely to be female, socioeconomically deprived, frontline workers, overweight or obese, and to have comorbidities (appendix table S11). Ongoing symptom severity also appeared to be linked to the severity of the initial infection: the proportion of participants who were either very unwell or hospitalised with their initial infection increased from 33% in the mild class to 60% in the severe class (appendix table S11); notably, however, 40% of participants in the severe class had an initial infection of mild-to-moderate severity (appendix table S11).

**Figure 1:**
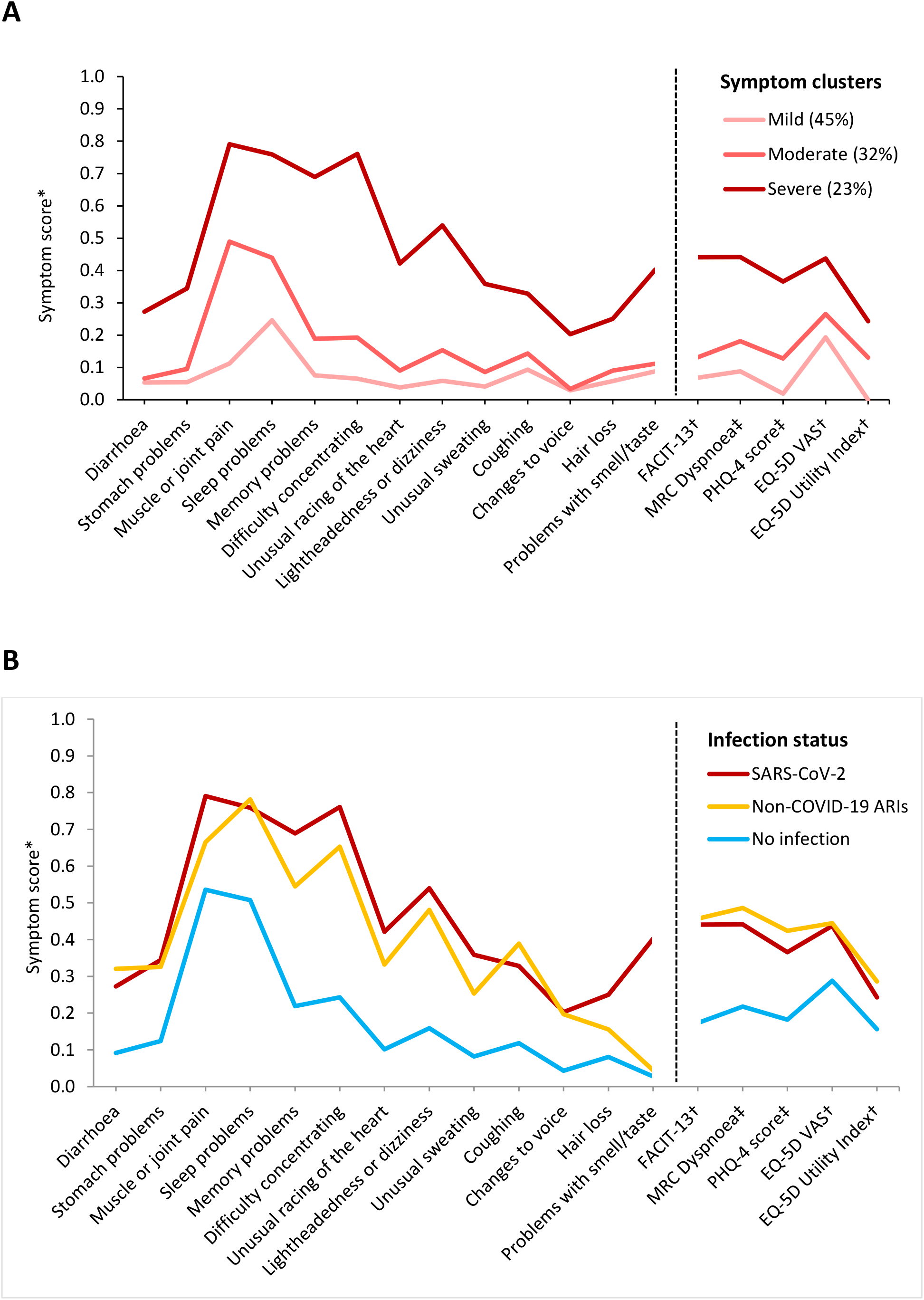
Symptom profiles among all participant with previous SARS-CoV-2 infection (A) and among participants with the most severe symptoms, by infection status (B) Figure shows the conditional probability (left of the dashed line) or mean severity (right of the dashed line) for all symptoms considered, adjusted for age and sex. Parallel lines between the classes indicate a fairly even increase in the probability or severity of the symptoms considered; disproportionate changes in symptom probability or severity is shown by deviations from the parallel. (A) Includes all participants with previous SARS-CoV-2. Displayed probabilities or scores are shown in appendix table S9. (B) Symptom profiles for participants with the most severe symptoms from the three separate latent class analyses are overlaid. MRC =Medical Research Council. VAS=visual analogue scale. PHQ=Patient Health Questionnaire. *Symptom score represents conditional probability for binary variables and mean severity for ordinal and continuous variables. †Continuous variables have been reversed to aid with interpretation, so that higher values indicate worse severity or health state. ‡Ordinal variable.

Finally, as ongoing symptom severity increased, participants were more likely to have reported having long COVID in the current questionnaire, ranging from 6.3% in the mild class to nearly half of participants in the severe class (appendix table S11). However, even in the severe class, many participants did not report themselves as having long COVID (appendix table S11).

Our final model for non-COVID-19 ARIs was also a three-class model representing different levels of severity (40% mild, 38% moderate, and 22% severe). This model had an entropy of 0.950 and showed strong distinction between classes (appendix table S9). To enable visual comparisons between participants by infection status, we additionally carried out an LCA in participants with no infection, resulting in a two-class model representing two levels of symptom severity (56% mild, 44% severe), and overlaid the symptom profiles of the severe class from each LCA (figure 1B). In this comparison, participants with non-COVID-19 ARI or SARS-CoV-2 infection showed a higher probability of reporting almost all symptoms than participants with no infection, particularly cognitive problems (figure 1B). However, symptom profiles seem to differ by ARI among those most severely affected: participants with SARS-CoV-2 infection showed a higher probability of reporting memory problems, difficulty concentrating, hair loss, and problems with sense of taste or smell compared with non-COVID-19 ARI, and slightly lower severity of dyspnoea and probability of coughing (figure 1B). Notably, a tenth of participants with previous non-COVID-19 ARI and severe symptoms attributed those symptoms to long COVID (appendix table S12). When comparing participant characteristics across the three groups with the most severe symptoms, participants with no reported infection were more likely be older than those with previous respiratory infections and presented with fewer symptoms overall, whereas participants with non-COVID-19 ARIs had a far higher prevalence of heart disease than the other groups (appendix table S12). Given this finding, we did an exploratory analysis comparing the prevalence and severity of all symptoms between participants with SARS-CoV-2 infection and those with non-COVID-19 ARI, additionally adjusting for heart disease, but this did not materially affect the results (data not shown).

## Discussion

In this large, observational study, we found that previous SARS-CoV-2 infection was associated with increased prevalence and severity of a wide range of symptoms—covering gastrointestinal, neurological, musculoskeletal, and cardiopulmonary problems—as well as lower HRQoL, and that this increased symptom burden persisted more than 12 weeks after the acute infection. Participants with SARS-CoV-2 infection were more likely to report problems with taste or smell and hair loss than those with non-COVID-19 ARI, but we observed little difference in other symptoms or HRQoL measures. The lower burden of coughing, problems with taste or smell, dyspnoea, and fatigue among participants who had been infected with SARS-CoV-2 more than 12 weeks prior compared with those with more recent infections suggests these symptoms may be the first to show improvement; however, other symptoms considered showed little difference between remote and more recent infections. A severe initial SARS-CoV-2 infection—either requiring bedrest or hospitalisation—was found to be associated with a greater prevalence of ongoing symptoms, and reduction in HRQoL. Finally, as ongoing symptom severity increased, participants were more likely to report having long COVID, with nearly half of participants with severe symptoms reporting suspected long COVID.

The scale and fast spread of the COVID-19 pandemic, alongside a lower case–fatality ratio than previous coronavirus pandemics,^29^ has resulted in hundreds of millions of survivors globally over just a few years, focusing attention on their post-COVID-19 experience. While post-acute sequelae of other viral respiratory infections have been observed,^30^ they have not been as well characterised. Similar to our findings for previous SARS-CoV-2 infection, we observed increased burden of many symptoms among participants with previous non-COVID-19 ARI when compared with no infection. However, long-term symptom profiles differed slightly between SARS-CoV-2 infection and non-COVID-19 ARI, with the former showing greater increases in problems with taste or smell, hair loss, and possibly memory problems. Our findings suggest that there may be long-lasting health impacts from other respiratory infections that are going unrecognised. Our cohort of community infections will not represent those worst affected by long COVID, and so the elevated symptom burden seen for both SARS-CoV-2 infections and non-COVID-19 ARIs is likely to represent a milder phenotype of post-acute sequelae. Indeed, retrospective cohort studies have generally found worse post-acute outcomes among patients with SARS-CoV-2 compared with influenza^31, 32^ and other viral infections;^33^ these studies have largely been restricted to hospitalised patients or electronic health record data, thus representing people with either a severe acute infection or sufficiently severe post-acute symptoms to seek medical help. Our findings of post-acute sequelae in people with milder disease, longitudinal studies are needed to investigate the distinct pathogens responsible and trajectories of recovery.

We found increased risk of ongoing symptoms across different levels of acute disease severity, highlighting the wide reach of long COVID burden. While long COVID has been well documented in previously hospitalised patients,^34–36^ and risk of long COVID has been found to be associated with severe COVID-19,^3, 37^ an increasing number of studies have found long-term sequelae in people with mild or asymptomatic infections.^11, 38^ Importantly, less than a quarter of our participants with previous SARS-CoV-2 infection had ever reported long COVID—including half of participants with the most severe symptoms—suggesting that some people with ongoing symptoms may not be ascribing these symptoms to the infection, or may not consider their symptoms serious enough to qualify as long COVID. This could exacerbate under-reporting of the condition,^39^ likely impacting the healthcare resources made available to people with long-term sequelae. A lack of awareness of post-acute sequelae of other ARIs—or even the lack of a common term, like ‘long COVID’—is likely to contribute to under-reporting as well. Among participants with the most severe symptoms, 12% with previous non-COVID-19 ARI attributed their symptoms to long COVID compared with 48% with previous SARS-CoV-2 infection, despite their symptom profiles being largely similar, highlighting their need for an alternative diagnosis. Given the few diagnostic tests available, future research needs to focus on enabling diagnosis of long COVID and other post-acute sequelae, to ensure all people with ongoing symptoms can access the support they need.

While exploring long COVID phenotypes, we found that people with previous SARS-CoV-2 infections could largely be classified into three groups representing different overall severity of ongoing symptoms. The most severely affected were characterised by neurocognitive symptoms such as memory problems and difficulty concentrating: these symptoms distinguished them not only from participants with milder ongoing symptoms after SARS-CoV-2 infection, but also from the most severely affected participants with non-COVID-19 ARI. Neurological and cognitive symptoms in long COVID are well documented,^2, 40^ and some studies have identified neurological symptom clusters as a potential long COVID phenotype.^3, 41^ However, research so far into phenotypes has been inconclusive, with other studies instead identifying clusters reflecting lower or higher overall symptom severity.^2, 42–44^ While our findings support the importance of neurocognitive symptoms in long COVID, the clearest distinction between our participants was the overall severity of their symptoms, rather than physiological systems affected. Such severity-based clusters may offer less insight into the underlying mechanisms of long COVID, but it will nonetheless be important to evaluate whether trajectories of recovery differ between these severity groups, in order to better predict outcomes for patients and support them in their recovery.

Our study has several strengths. By comparing symptom burden across three groups, we can paint a clearer picture of how post-acute sequelae differ between SARS-CoV-2 and other ARIs, and how these symptoms differ from those experienced by uninfected population controls. The importance of a control population can be seen clearly when considering muscle and joint pain and sleep problems: while more frequent in people with previous ARI, these symptoms are also widespread in the uninfected population, and so may not be the most characteristic of previous ARIs. Our large sample includes a mix of asymptomatic, symptomatic, and hospitalised cases, allowing us to examine post-acute sequelae across all severities of the acute infection. We recorded the presence and severity of potential long COVID symptoms from all participants in our cohort, regardless of previous SARS-CoV-2 infection status or long COVID reports, which reduces potential reporting bias. This also enabled us to compare responses across all participants at the same point in the pandemic, effectively allowing us to adjust for baseline levels of symptoms such as fatigue and depression, which were likely elevated in the general population owing to the stress of the pandemic.^12^ Access to pre-vaccination antibody test data for more than half of our participants, both from COVIDENCE UK serological sub-studies^45^ and as reported by our participants, allowed us to more accurately identify previous infections and thus more robustly classify our controls.

This study also has some limitations. We focused on symptoms for each individual at a single timepoint, and thus could not map the change in each participant’s symptoms over time with repeated measures; however, our aim for this study was to provide a descriptive snapshot of the post-COVID-19 symptom burden, and longitudinal analyses are planned for future work. Additionally, we did not adjust for pre-COVID-19 symptom burden, as few participants had these data available, meaning that we cannot know whether participants with previous infections had a higher baseline symptom burden than those without. However, our inclusion of all participants with previous SARS-CoV-2 infection, rather than only symptomatic or severe COVID-19, reduces the risk of baseline differences, as many determinants of SARS-CoV-2 infection reflect lifestyle or behavioural factors rather than biological ones.^45, 46^ Additionally, sensitivity analyses adjusting for pre-infection general health did not substantially affect our results. We do not have details on the type of respiratory infections experienced by our participants reporting non-COVID-19 ARIs, as these are not routinely tested for in the community. However, we did require a negative SARS-CoV-2 swab test for non-COVID-19 ARIs, and owing to the timing of our study, the reported tests are likely to have been RT-PCR tests, reducing the risk of false negatives.^47^ Additionally, a COVIDENCE UK serological sub-study was carried out in December, 2020,^45^ meaning we had recent serology data for approximately 40% of our participants, helping to reduce misclassification. As COVIDENCE UK questionnaires are issued monthly, participants may have forgotten to report symptoms from minor ARIs that had resolved several weeks earlier. However, more than 60% of participants with ARIs reported being mildly unwell, suggesting that our findings have not been skewed towards people with the most severe infections. The final models chosen for the clustering analysis had evidence of residual covariance between the included variables (appendix p 15), suggesting that the latent class assignments did not fully explain the symptom distribution between participants; nonetheless, the models showed strong distinction between classes, and our findings of clusters defined by symptom severity mirror those of several other studies,^42–44^ lending strength to our results. Our findings are restricted to unvaccinated patients infected with either the wild-type or alpha strains of SARS-CoV-2. However, 29% of people in the UK reporting long COVID were infected with the wild-type strain,^5^ which dominated before the vaccination rollout in the UK, showing that our participants represent an important and substantial subgroup with long-lasting symptoms. Many of our symptoms were recorded as binary measures, requiring participants to classify themselves as either having them or not; more use of scales, such as MRC Dyspnoea or FACIT-13, would have added further nuance to our findings. Finally, women, older age groups, and White ethnicity are over-represented in our study, potentially limiting the generalisability of our results.

The COVID-19 pandemic has cast a much-needed spotlight on post-acute infection syndromes, highlighting the need for improved understanding, diagnosis, and treatment of these conditions. While the high symptom burden we observed in participants with previous SARS-CoV-2 infection illustrates the extensive reach of long COVID, the similar burden observed among people with previous non-COVID-19 ARI suggests that the lasting impacts of these infections may be underestimated. As research into long COVID continues, we must take the opportunity to investigate and consider the post-acute burden of other ARIs, to ensure all people with post-acute sequelae can access the treatment and care they deserve.

## Contributors

GV, ARM, and PEP conceived the analysis. GV and MT contributed to data management, and have directly accessed and verified the data. Statistical analyses were done by GV, with input from PEP, ARM, MT, SOS, and JB. GV wrote the first draft of the report. All authors critically revised the manuscript for intellectually important content. All authors provided critical conceptual input, interpreted the data analysis, and read and approved the final draft. GV and ARM had final responsibility for the decision to submit the manuscript for publication.

## Data sharing statement

De-identified participant data will be made available upon reasonable request to the corresponding author.

## Acknowledgments

COVIDENCE UK has received support from Barts Charity (MGU0459, MGU0466), Pharma Nord, the Fischer Family Foundation, DSM Nutritional Products, the Exilarch’s Foundation, the Karl R Pfleger Foundation, the AIM Foundation, Synergy Biologics, Cytoplan, the UK National Institute for Health and Care Research Clinical Research Network (52255; 52257), the Health Data Research UK BREATHE Hub, the UK Research and Innovation Industrial Strategy Challenge Fund (MC_PC_19004), Thornton & Ross, Warburtons, Matthew Isaacs (personal donation), Barbara Boucher (personal donation), and Hyphens Pharma. MT was supported by Barts Charity (MGU0570). The views expressed are those of the authors and not necessarily those of the funders. We thank all participants of COVIDENCE UK, and the following organisations who supported study recruitment: Asthma UK/British Lung Foundation, the British Heart Foundation, the British Obesity Society, Cancer Research UK, Diabetes UK, Future Publishing, Kidney Care UK, Kidney Wales, Mumsnet, the National Kidney Federation, the National Rheumatoid Arthritis Society, the North West London Health Research Register (DISCOVER), Primary Immunodeficiency UK, the Race Equality Foundation, SWM Health, the Terence Higgins Trust, and Vasculitis UK.

