## Appendix for "Long-term symptom profiles after COVID-19 *vs* other acute respiratory infections: a population-based observational study (COVIDENCE UK)"

**Supplementary appendix**

### ***Figure S1:* Study flow diagram**

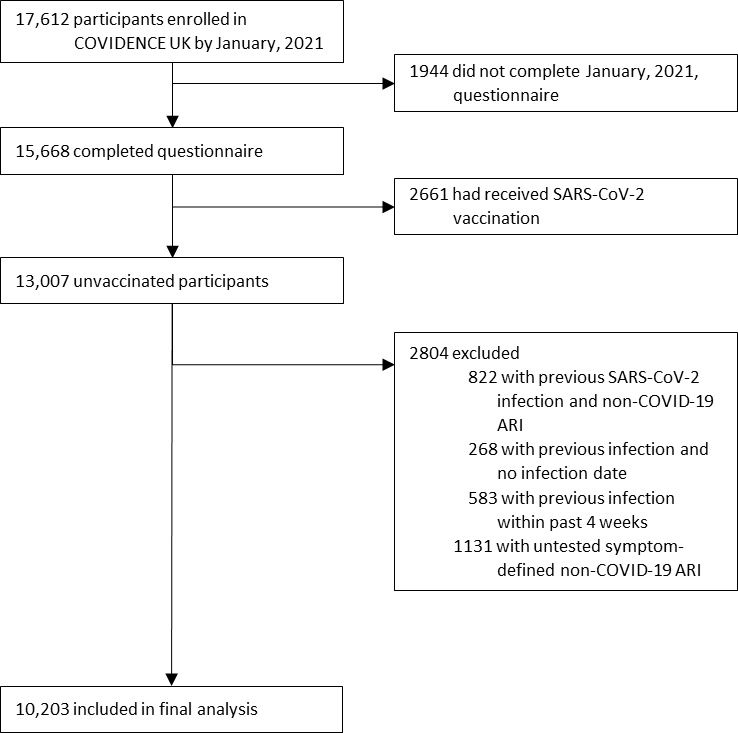

### **Regression analysis**

Our outcome measures were a mix of binary (eg, presence or absence of symptoms), ordinal (eg, symptoms measured on a discrete scale), and continuous bounded (eg, symptom scores ranging from 0 to 100). We therefore used different regression techniques to model each of these outcomes.

|  | **Description** |
| --- | --- |
| Logistic regression | - Measures the probability of an event occurring - Output can be presented as a single odds ratio |
| Ordinal logistic regression | - Extension of the logistic regression model to outcomes with more than two ordered categories - Output can be presented as a single odds ratio, which represents the difference between *any two sequential categories* - Relies on the proportional odds assumption that the effect coefficient $\beta$ of a given explanatory variable is the same across all levels of the outcome (in practical terms, that the odds ratio between any two sequential categories will be the same) |
| Partial proportional odds model | - Can be used when the proportional odds assumption is not satisfied - Sits between ordinal regression and the generalised ordered logit model - Allows some—but not necessarily all—of the $\beta$ coefficients to vary across different levels of the outcome variable, depending on whether they violate the proportional odds assumption - A more parsimonious approach (ie, requires fewer parameters) than removing proportional odds constraints from all variables in the model |
| Fractional regression | - An alternative to linear regression for bounded variables, defined on the unit interval [0,1]; in such cases, linear regression may not be appropriate, and can yield problems when predictions for the response variables approach the interval boundaries - Useful when boundary values are common (eg, when reporting no symptoms) - Estimates are very similar to linear regression in the cases that linear regression does not present a problem |

*Assessment of proportional odds assumption*

The proportional odds assumption for ordinal variables was assessed using the Brant test^1^ (where this could not be run, a likelihood ratio test between unconstrained and constrained generalised ordered logit models was used). Models with significant deviations were refitted using a partial proportional odds model.^2^ As deviations are common with larger sample sizes, model fit was assessed by comparing Lacy’s adjusted ordinal explained variation measure, $R_{O}^{2}$adj,^3^ between the ordered logistic regression model and the partial proportional odds model. As partial proportional odds models require a greater number of parameters to be estimated, we only examined those models with a greater than 5% increase in $R_{O}^{2}$adj (note: *not* percentage-point increase) to preserve model parsimony.^3^

Eight partial proportional odds models presented with more than a 5% increase in $R_{O}^{2}$adj when compared with their respective ordered logistic regression models: eight for PHQ-4 and two for EQ5D Activities. The maximum increase in $R_{O}^{2}$adj was 10%. The covariate of interest (ie, infection status, infection timing, or infection severity) did not violate the proportional odds assumption in any of the models considered, and so all models produced one odds ratio for a change in level of the dependent variable. Odds ratios and 95% CIs for the covariate of interest were identical for four of the models considered. Where estimates differed, we used findings from the partial proportional odds model. The six models with different findings are listed below.

|  | **Change in** $\boldsymbol{R}_{\boldsymbol{O}}^{\boldsymbol{2}}$**adj** | **OR (95% CI)** | |
| --- | --- | --- | --- |
|  |  | Ordered logistic regression | Partial proportional odds |
| **Main analysis** | | | |
| COVID-19 severity (EQ-5D Activities) | 7% | 1.32 (0.56–3.12) | 1.33 (0.56–3.14) |
|  |  | 1.85 (0.82–4.15) | 1.87 (0.83–4.22) |
|  |  | 2.45 (1.14–5.27) | 2.32 (1.07–5.01) |
|  |  | 10.16 (3.88–26.56) | 12.58 (4.60–34.41) |
| **Sensitivity analysis 1** | | | |
| COVID-19 *vs* no infection (PHQ-4) | 7% | 1.22 (0.99–1.49) | 1.22 (1.00–1.49) |
| COVID-19 severity (EQ-5D Activities) | 10% | 0.35 (0.07–1.83) | 0.35 (0.07–1.81) |
|  |  | 1.74 (0.59–5.10) | 1.77 (0.60–5.23) |
|  |  | 3.64 (1.43–9.28) | 3.37 (1.32–8.62) |
|  |  | 11.63 (4.07–33.24) | 14.79 (4.94–44.27) |
| **Sensitivity analysis 2** | | | |
| COVID-19 *vs* no infection (PHQ-4) | 8% | 1.35 (1.02–1.78) | 1.34 (1.02–1.77) |
| COVID-19 >12 weeks prior *vs* no infection (PHQ-4) | 8% | 0.99 (0.63–1.55) | 0.98 (0.63–1.53) |
| Non-COVID-19 ARI *vs* no infection (PHQ-4) | 8% | 1.30 (1.05–1.61) | 1.30 (1.05–1.60) |

OR=odds ratio. PHQ=Patient Health Questionnaire.

### ***Table S1:* Comparison between findings of ordered logistic regression models and partial proportional odds models**

### ***Figure S2:* Symptom prevalence by infection status**

By infection status

Percentage of participants*

|  | No infection | Non-COVID-19 ARI | SARS-CoV-2 infection |
| --- | --- | --- | --- |
| Coughing | 8.1 | 25.7 | 15.8 |
| Problems with sleeping | 33.8 | 51 | 40.2 |
| Memory problems | 12.2 | 22.8 | 20.8 |
| Difficulty concentrating | 13 | 28.6 | 22.8 |
| Pains in muscles or joints | 30.2 | 41.7 | 35.9 |
| Problems with sense of smell / taste | 1.9 | 4.7 | 12.5 |
| Diarrhoea | 5.3 | 15.3 | 10.2 |
| Stomach (abdominal pains) | 6.8 | 17.7 | 10.8 |
| Changes to voice | 2.3 | 9.1 | 6.8 |
| Hair loss | 5 | 8.3 | 8.5 |
| Unusual racing of the heart | 5.6 | 15.6 | 11.6 |
| Lightheadedness or dizziness | 8.8 | 21.3 | 16.6 |
| Unusual sweating | 4.7 | 13.3 | 9.9 |
| MRC Dyspnoea 1 | 74.7 | 60 | 67.1 |
| MRC Dyspnoea 2 | 22.1 | 32.2 | 26.3 |
| MRC Dyspnoea 3 | 2.4 | 5.6 | 5 |
| MRC Dyspnoea 4 | 0.6 | 1.6 | 1.2 |
| MRC Dyspnoea 5 | 0.1 | 0.6 | 0.4 |
| PHQ-4 Normal | 81 | 65 | 74.7 |
| PHQ-4 Mild | 14.2 | 24.8 | 18.7 |
| PHQ-4 Moderate | 3.2 | 6.1 | 4.3 |
| PHQ-4 Severe | 1.7 | 4.1 | 2.3 |
| FACIT-13 (reversed)* | 8 | 13 | 10 |
| Activities No Problems | 92.1 | 81.2 | 85.1 |
| Activities Some Problems | 7.5 | 17.2 | 13.8 |
| Activities Unable | 0.4 | 1.6 | 1 |
| Anxiety or depression None | 73.2 | 61.6 | 66.8 |
| Anxiety or depression Moderate | 25.7 | 36 | 31.6 |
| Anxiety or depression Extreme | 1.1 | 2.4 | 1.6 |
| Pain or discomfort None | 76 | 61.5 | 70.4 |
| Pain or discomfort Moderate | 22.7 | 34.6 | 27.6 |
| Pain or discomfort Extreme | 1.4 | 3.8 | 2 |
| Self-care No Problems | 97.7 | 94.4 | 96.5 |
| Self-care Some Problems | 2.2 | 5.4 | 3.4 |
| Self-care Unable | 0.1 | 0.2 | 0.1 |
| Mobility No Problems | 90.6 | 84.8 | 88.8 |
| Mobility Some Problems | 9.3 | 14.8 | 11 |
| Mobility Unable | 0.1 | 0.5 | 0.2 |
| EQ5D VAS* | 83 | 75 | 80 |

VAS=visual analogue scale. *Median scores are presented for continuous variables. FACIT-13 has been rescaled and reversed, so that 0 represents no fatigue and 100 represents maximum fatigue. For EQ-5D VAS, 0 represents worst possible health and 100 represents perfect health.

### ***Figure S3:* Symptom prevalence by infection timing**

VAS=visual analogue scale. *Median scores are presented for continuous variables. FACIT-13 has been rescaled and reversed, so that 0 represents no fatigue and 100 represents maximum fatigue. For EQ-5D VAS, 0 represents worst possible health and 100 represents perfect health.

By SARS-CoV-2 infection timing

Percentage of participants*

|  | 4–12 weeks | >12 weeks |
| --- | --- | --- |
| Coughing | 28.7 | 14.6 |
| Problems with sleeping | 45.8 | 43.4 |
| Memory problems | 25.3 | 24.4 |
| Difficulty concentrating | 30.2 | 25.9 |
| Pains in muscles or joints | 35.8 | 37.8 |
| Problems with sense of smell / taste | 26.6 | 11.9 |
| Diarrhoea | 19.1 | 9.4 |
| Stomach (abdominal pains) | 9.6 | 12.4 |
| Changes to voice | 11.7 | 5.7 |
| Hair loss | 8.5 | 10.7 |
| Unusual racing of the heart | 13.8 | 13.2 |
| Lightheadedness or dizziness | 21.3 | 19 |
| Unusual sweating | 15.8 | 11.7 |
| MRC Dyspnoea 1 | 57.3 | 68.7 |
| MRC Dyspnoea 2 | 30.2 | 25.1 |
| MRC Dyspnoea 3 | 10.4 | 5.1 |
| MRC Dyspnoea 4 | 0 | 1.2 |
| MRC Dyspnoea 5 | 2.1 | 0 |
| PHQ-4 Normal | 66.3 | 72.9 |
| PHQ-4 Mild | 28.4 | 19.1 |
| PHQ-4 Moderate | 4.2 | 5.1 |
| PHQ-4 Severe | 1.1 | 2.9 |
| FACIT-13 (reversed)* | 15 | 12 |
| Activities No Problems | 79.2 | 84.1 |
| Activities Some Problems | 17.7 | 14.6 |
| Activities Unable | 3.1 | 1.3 |
| Anxiety or depression None | 68.8 | 64.7 |
| Anxiety or depression Moderate | 30.2 | 33.3 |
| Anxiety or depression Extreme | 1 | 2 |
| Pain or discomfort None | 66.7 | 67.3 |
| Pain or discomfort Moderate | 32.3 | 30.2 |
| Pain or discomfort Extreme | 1 | 2.5 |
| Self-care No Problems | 99 | 95.4 |
| Self-care Some Problems | 0 | 4.6 |
| Self-care Unable | 1 | 0 |
| Mobility No Problems | 88.5 | 88.2 |
| Mobility Some Problems | 10.4 | 11.5 |
| Mobility Unable | 1 | 0.2 |
| EQ5D VAS* | 72 | 79 |

### ***Figure S4:* Symptom prevalence by infection severity**

By SARS-CoV-2 infection severity

Percentage of participants*

|  | Asymptomatic | Mild | Moderate | Very unwell | Hospitalised |
| --- | --- | --- | --- | --- | --- |
| Coughing | 5.9 | 16.1 | 24.7 | 28.9 | 20.6 |
| Problems with sleeping | 38.3 | 34.9 | 43.5 | 51.9 | 64.7 |
| Memory problems | 10.5 | 14 | 20.5 | 31.6 | 58.8 |
| Difficulty concentrating | 11 | 15.1 | 25.9 | 34.9 | 64.7 |
| Pains in muscles or joints | 31.2 | 34.4 | 41.1 | 44.1 | 58.8 |
| Problems with sense of smell / taste | 5.2 | 21.5 | 18.9 | 23.2 | 20.6 |
| Diarrhoea | 5.2 | 9.1 | 18.4 | 17.1 | 26.5 |
| Stomach (abdominal pains) | 6.5 | 7.5 | 10.8 | 17.5 | 23.5 |
| Changes to voice | 3.3 | 7 | 8.1 | 15.7 | 14.7 |
| Hair loss | 6.5 | 4.8 | 4.3 | 13.3 | 8.8 |
| Unusual racing of the heart | 10.4 | 5.9 | 10.8 | 15.6 | 38.2 |
| Lightheadedness or dizziness | 12.3 | 11.3 | 22.7 | 28.9 | 41.2 |
| Unusual sweating | 5.8 | 8.1 | 9.2 | 20.4 | 14.7 |
| MRC Dyspnoea 1 | 71.6 | 71 | 62.4 | 57.1 | 29.4 |
| MRC Dyspnoea 2 | 26.5 | 25.8 | 31.2 | 26.4 | 38.2 |
| MRC Dyspnoea 3 | 1.3 | 2.2 | 6.5 | 11.3 | 26.5 |
| MRC Dyspnoea 4 | 0.6 | 1.1 | 0 | 2.8 | 5.9 |
| MRC Dyspnoea 5 | 0 | 0 | 0 | 2.4 | 0 |
| PHQ-4 Normal | 77.4 | 77.4 | 72.3 | 65.7 | 52.9 |
| PHQ-4 Mild | 18.1 | 18.3 | 19.6 | 23.8 | 23.5 |
| PHQ-4 Moderate | 1.9 | 2.2 | 7.1 | 7.6 | 11.8 |
| PHQ-4 Severe | 2.6 | 2.2 | 1.1 | 2.9 | 11.8 |
| FACIT-13 (reversed)* | 8 | 10 | 13 | 17 | 40 |
| Activities No Problems | 91 | 89.8 | 80.1 | 76.4 | 38.2 |
| Activities Some Problems | 9 | 10.2 | 19.9 | 19.8 | 61.8 |
| Activities Unable | 0 | 0 | 0 | 3.8 | 0 |
| Anxiety or depression None | 65.2 | 69.4 | 69.7 | 59 | 47.1 |
| Anxiety or depression Moderate | 34.2 | 29.6 | 27 | 38.7 | 38.2 |
| Anxiety or depression Extreme | 0.6 | 1.1 | 3.2 | 2.4 | 14.7 |
| Pain or discomfort None | 75.5 | 74.2 | 66.1 | 63 | 52.9 |
| Pain or discomfort Moderate | 23.2 | 25.3 | 31.2 | 34.1 | 32.4 |
| Pain or discomfort Extreme | 1.3 | 0.5 | 2.7 | 2.8 | 14.7 |
| Self-care No Problems | 98.7 | 97.8 | 96.2 | 94.3 | 73.5 |
| Self-care Some Problems | 1.3 | 2.2 | 3.8 | 5.2 | 26.5 |
| Self-care Unable | 0 | 0 | 0 | 0.5 | 0 |
| Mobility No Problems | 90.9 | 89.2 | 88.2 | 84.4 | 60.6 |
| Mobility Some Problems | 9.1 | 10.8 | 11.8 | 15.1 | 39.4 |
| Mobility Unable | 0 | 0 | 0 | 0.5 | 0 |
| EQ5D VAS* | 82 | 80 | 77 | 70 | 58 |

VAS=visual analogue scale. *Median scores are presented for continuous variables. FACIT-13 has been rescaled and reversed, so that 0 represents no fatigue and 100 represents maximum fatigue. For EQ-5D VAS, 0 represents worst possible health and 100 represents perfect health.

|  | **FACIT-13 score (reversed)** | **EQ-5D VAS** |
| --- | --- | --- |
| **By infection status (table 1)** |  |  |
| Previous SARS-CoV-2 infection *vs* no infection | 0.32 (0.24 to 0.39) | -0.11 (-0.16 to -0.06) |
| SARS-CoV-2 infection >12 weeks prior *vs* no infection | 0.28 (0.20 to 0.36) | -0.09 (-0.15 to -0.04) |
| Previous SARS-CoV-2 infection *vs* non-COVID-19 ARI | -0.02 (-0.15 to 0.10) | 0.01 (-0.09 to 0.10) |
| Previous non-COVID-19 ARI *vs* no infection | 0.33 (0.22 to 0.44) | -0.09 (-0.17 to 0.00) |
| **By SARS-CoV-2 infection timing (table 2)** |  |  |
| SARS-CoV-2 infection >12 weeks prior *vs* 4–12 weeks prior | -0.29 (-0.49 to -0.08) | 0.11 (-0.03 to 0.26) |
| **By SARS-CoV-2 infection severity (table 3)** |  |  |
| Mildly unwell *vs* asymptomatic | 0.09 (-0.20 to 0.39) | -0.16 (-0.39 to 0.08) |
| Moderately unwell *vs* asymptomatic | 0.11 (-0.18 to 0.40) | 0.02 (-0.20 to 0.25) |
| Very unwell *vs* asymptomatic | 0.43 (0.15 to 0.72) | -0.12 (-0.34 to 0.09) |
| Hospitalised *vs* asymptomatic | 1.21 (0.76 to 1.65) | -0.38 (-0.70 to -0.06) |

Data are estimate (95% CI). ARI=acute respiratory infection. VAS=visual analogue scale.

### ***Table S2:* Raw coefficients for changes in FACIT-13 score and EQ-5D VAS**

|  | **Previous SARS-CoV-2 infection *vs* no infection** | | **SARS-CoV-2 infection >12 weeks prior *vs* no infection** | | **Previous SARS-CoV-2 infection *vs* previous non-COVID-19 ARI** | |
| --- | --- | --- | --- | --- | --- | --- |
|  | Estimate (95% CI)* | p value† | Estimate (95% CI)* | p value‡ | Estimate (95% CI)* | p value§ |
| Coughing | 2.05 (1.61–2.61) | <0.001 | 1.48 (1.09–2.00) | 0.011 | 0.80 (0.57–1.11) | 0.179 |
| Problems with sleep | 1.25 (1.04–1.49) | 0.015 | 1.22 (1.00–1.50) | 0.050 | 0.76 (0.58–1.00) | 0.051 |
| Memory problems | 2.11 (1.71–2.61) | <0.001 | 2.01 (1.58–2.55) | <0.001 | 1.48 (1.07–2.04) | 0.017 |
| Difficulty concentrating | 1.83 (1.48–2.27) | <0.001 | 1.79 (1.41–2.28) | <0.001 | 1.29 (0.95–1.76) | 0.107 |
| Pains in muscles or joints | 1.29 (1.07–1.56) | 0.007 | 1.25 (1.01–1.54) | 0.042 | 1.06 (0.81–1.40) | 0.667 |
| Problems with sense of smell / taste | 11.67 (8.93–15.24) | <0.001 | 10.25 (7.60–13.82) | <0.001 | 11.63 (6.30–21.47) | <0.001 |
| Diarrhoea | 1.57 (1.16–2.13) | 0.003 | 1.39 (0.98–1.98) | 0.067 | 0.87 (0.58–1.31) | 0.506 |
| Stomach (abdominal pains) | 1.62 (1.23–2.12) | <0.001 | 1.61 (1.18–2.18) | 0.002 | 0.77 (0.53–1.12) | 0.174 |
| Changes to voice | 3.20 (2.25–4.54) | <0.001 | 3.06 (2.07–4.52) | <0.001 | 1.04 (0.64–1.68) | 0.873 |
| Hair loss | 2.03 (1.51–2.72) | <0.001 | 2.12 (1.54–2.93) | <0.001 | 2.14 (1.31–3.49) | 0.002 |
| Unusual racing of the heart | 2.47 (1.91–3.20) | <0.001 | 2.45 (1.83–3.27) | <0.001 | 1.43 (0.98–2.10) | 0.066 |
| Lightheadedness or dizziness | 2.09 (1.66–2.64) | <0.001 | 1.99 (1.53–2.59) | <0.001 | 1.32 (0.93–1.88) | 0.114 |
| Unusual sweating | 2.39 (1.81–3.15) | <0.001 | 2.23 (1.63–3.07) | <0.001 | 1.45 (0.96–2.20) | 0.077 |
| MRC Dyspnoea\|\| | 1.60 (1.32–1.94) | <0.001 | 1.40 (1.13–1.75) | 0.003 | 1.21 (0.91–1.60) | 0.190 |
| PHQ-4 grade\|\| | 1.22 (1.00–1.49) | 0.058 | 1.07 (0.89–1.30) | 0.457 | 0.85 (0.64–1.14) | 0.281 |
| FACIT-13 score (reversed)¶ | 3.8% (2.5–5.1) | <0.001 | 2.9% (1.4–4.3) | <0.001 | 0.5% (-1.8–2.7) | 0.693 |
| EQ-5D Activities\|\| | 2.27 (1.76–2.94) | <0.001 | 2.11 (1.58–2.82) | <0.001 | 1.16 (0.81–1.66) | 0.409 |
| EQ-5D Pain\|\| | 1.35 (1.11–1.64) | 0.003 | 1.32 (1.06–1.64) | 0.015 | 0.95 (0.72–1.27) | 0.754 |
| EQ-5D Self-care (two groups) | 0.90 (0.53–1.53) | 0.699 | 1.03 (0.59–1.79) | 0.912 | 1.04 (0.49–2.22) | 0.912 |
| EQ-5D Mobility (two groups) | 1.10 (0.82–1.49) | 0.516 | 1.02 (0.73–1.43) | 0.917 | 0.91 (0.60–1.38) | 0.644 |
| EQ-5D VAS¶ | -2.1% (-3.5 to -0.7) | 0.004 | -1.5% (-3.1 to 0.0) | 0.055 | 0.5% (-1.8 to 2.7) | 0.692 |

Analysis is done in 579 participants with previous SARS-CoV-2 infection (502 symptomatic), 472 participants with previous non-COVID-19 ARI, and 8388 participants with no infection. ARI=acute respiratory infection. PHQ=Patient Health Questionnaire. MRC=Medical Research Council. VAS=visual analogue scale. *Estimates are odds ratios for binary and ordinal outcomes, and predicted percentage point changes for continuous outcomes. †After adjustment for multiple testing, p<0.04 is the threshold for statistical significance. ‡After adjustment for multiple testing, p<0.03 is the threshold for statistical significance. § After adjustment for multiple testing, p<0.005 is the threshold for statistical significance. ||Ordinal outcome. ¶Continuous outcome.

### ***Table S3:* Symptom comparisons among participants with test-confirmed previous SARS-CoV-2 infection *vs* non-COVID-19 ARI or no infection**

|  | **SARS-CoV-2 infection >12 weeks prior *vs* <4 weeks prior** | |
| --- | --- | --- |
|  | Estimate* (95% CI) | p value† |
| Coughing | 0.31 (0.19–0.52) | <0.001 |
| Problems with sleep | 0.97 (0.64–1.47) | 0.882 |
| Memory problems | 0.84 (0.52–1.36) | 0.472 |
| Difficulty concentrating | 0.89 (0.55–1.44) | 0.640 |
| Pains in muscles or joints | 0.97 (0.64–1.49) | 0.897 |
| Problems with sense of smell / taste | 0.62 (0.38–1.01) | 0.054 |
| Diarrhoea | 0.56 (0.29–1.11) | 0.097 |
| Stomach (abdominal pains) | 0.99 (0.52–1.88) | 0.975 |
| Changes to voice | 0.72 (0.33–1.60) | 0.422 |
| Hair loss | 1.16 (0.56–2.40) | 0.679 |
| Unusual racing of the heart | 0.98 (0.55–1.75) | 0.943 |
| Lightheadedness or dizziness | 0.88 (0.52–1.47) | 0.617 |
| Unusual sweating | 0.68 (0.36–1.25) | 0.214 |
| MRC Dyspnoea‡ | 0.61 (0.40–0.94) | 0.025 |
| PHQ-4 grade‡ | 0.82 (0.57–1.18) | 0.284 |
| FACIT-13 score (reversed)§ | -4.3% (-8.0 to -0.5) | 0.028 |
| EQ-5D Activities‡ | 0.85 (0.49–1.49) | 0.914 |
| EQ-5D Pain‡ | 1.02 (0.66–1.60) | 0.523 |
| EQ-5D Self-care (two groups) | 3.00 (0.34–26.55) | 0.502 |
| EQ-5D Mobility (two groups) | 0.80 (0.41–1.55) | 0.028 |
| EQ-5D VAS§ | 2.0% (-1.5 to 5.5) | 0.254 |

PHQ=Patient Health Questionnaire. MRC=Medical Research Council. VAS=visual analogue scale. *Estimates are odds ratios for binary and ordinal outcomes, and predicted percentage point changes for continuous outcomes. †After adjustment for multiple testing, p<0.002 is the threshold for statistical significance. ‡Ordinal outcome. §Continuous outcome.

### ***Table S4:* Symptom associations by time since infection, among participants with test-confirmed previous SARS-CoV-2 infection**

|  | **Mildly unwell *vs*  asymptomatic** | | **Moderately unwell *vs*  asymptomatic** | | **Very unwell *vs*  asymptomatic** | | **Hospitalised *vs*  asymptomatic** | |
| --- | --- | --- | --- | --- | --- | --- | --- | --- |
|  | Estimate* (95% CI) | p value† | Estimate* (95% CI) | p value† | Estimate* (95% CI) | p value‡ | Estimate* (95% CI) | p value§ |
| Coughing | 5.60 (1.44–21.80) | 0.013 | 3.91 (0.99–15.40) | 0.051 | 9.75 (2.76–34.46) | <0.001 | 6.71 (1.57–28.70) | 0.010 |
| Problems with sleep | 0.51 (0.24–1.11) | 0.091 | 0.90 (0.43–1.87) | 0.783 | 1.07 (0.56–2.07) | 0.834 | 1.63 (0.66–4.06) | 0.294 |
| Memory problems | 1.27 (0.44–3.66) | 0.653 | 2.24 (0.87–5.78) | 0.094 | 3.77 (1.63–8.74) | 0.002 | 10.11 (3.65–28.01) | <0.001 |
| Difficulty concentrating | 1.16 (0.35–3.79) | 0.810 | 2.63 (0.94–7.31) | 0.065 | 5.44 (2.19–13.50) | <0.001 | 16.65 (5.55–49.95) | <0.001 |
| Pains in muscles or joints | 0.81 (0.37–1.78) | 0.599 | 0.90 (0.42–1.91) | 0.782 | 1.59 (0.82–3.07) | 0.171 | 3.61 (1.48–8.81) | 0.005 |
| Problems with sense of smell / taste | 4.12 (1.34–12.67) | 0.013 | 5.59 (1.89–16.54) | 0.002 | 6.13 (2.21–17.04) | <0.001 | 6.75 (2.04–22.30) | 0.002 |
| Diarrhoea | 1.84 (0.28–12.03) | 0.525 | 6.64 (1.33–33.19) | 0.021 | 4.37 (0.92–20.87) | 0.064 | 13.00 (2.42–69.95) | 0.003 |
| Stomach (abdominal pains) | 1.36 (0.36–5.19) | 0.649 | 1.74 (0.51–5.99) | 0.378 | 1.93 (0.63–5.86) | 0.248 | 6.75 (1.93–23.68) | 0.003 |
| Changes to voice | 2.38 (0.47–12.19) | 0.297 | 1.94 (0.38–9.83) | 0.421 | 3.90 (0.96–15.92) | 0.058 | 3.54 (0.67–18.78) | 0.138 |
| Hair loss | 0.98 (0.28–3.49) | 0.975 | 0.89 (0.25–3.16) | 0.852 | 1.57 (0.56–4.41) | 0.388 | 2.49 (0.68–9.03) | 0.166 |
| Unusual racing of the heart | 0.84 (0.25–2.86) | 0.776 | 0.75 (0.23–2.45) | 0.636 | 1.62 (0.63–4.15) | 0.317 | 5.65 (1.88–16.97) | 0.002 |
| Lightheadedness or dizziness | 0.58 (0.16–2.04) | 0.394 | 2.28 (0.88–5.91) | 0.090 | 2.26 (0.95–5.36) | 0.065 | 4.01 (1.45–11.06) | 0.007 |
| Unusual sweating | 2.16 (0.58–8.14) | 0.253 | 0.72 (0.15–3.40) | 0.682 | 6.99 (2.34–20.86) | <0.001 | 1.20 (0.26–5.57) | 0.815 |
| MRC Dyspnoea\|\| | 1.68 (0.75–3.77) | 0.211 | 1.79 (0.82–3.89) | 0.143 | 2.30 (1.14–4.66) | 0.021 | 7.05 (2.91–17.07) | <0.001 |
| PHQ-4 grade\|\| | 0.83 (0.34–2.02) | 0.689 | 1.10 (0.47–2.56) | 0.823 | 1.38 (0.66–2.89) | 0.390 | 3.98 (1.58–9.99) | 0.003 |
| FACIT-13 score (reversed)¶ | -0.3% (-5.3 to 4.6) | 0.891 | 0.5% (-4.5 to 5.5) | 0.840 | 8.4% (2.9–21.8) | 0.003 | 21.8% (12.8–30.8) | <0.001 |
| EQ-5D Activities\|\| | 0.35 (0.07–1.81) | 0.210 | 1.77 (0.60–5.23) | 0.302 | 3.37 (1.32–8.62) | 0.011 | 14.79 (4.94–44.27) | <0.001 |
| EQ-5D Pain\|\| | 0.94 (0.42–2.08) | 0.873 | 0.78 (0.36–1.71) | 0.536 | 0.97 (0.49–1.92) | 0.921 | 3.11 (1.26–7.68) | 0.014 |
| EQ-5D Mobility (two groups) | 0.79 (0.17–3.60) | 0.760 | 1.13 (0.29–4.31) | 0.861 | 1.83 (0.59–5.65) | 0.292 | 8.06 (2.43–26.80) | <0.001 |
| EQ-5D VAS¶ | 1.4% (-4.8 to 7.5) | 0.661 | 0.6% (-5.2 to 6.3) | 0.850 | -3.4% (-8.9 to 2.0) | 0.219 | -7.8% (-15.1 to -0.6) | 0.034 |

Differences in EQ-5D Self Care were not estimable due to small numbers. PHQ=Patient Health Questionnaire. MRC=Medical Research Council. VAS=visual analogue scale. *Estimates are odds ratios for binary and ordinal outcomes, and predicted percentage point changes for continuous outcomes. †After adjustment for multiple testing, p<0.002 is the threshold for statistical significance. ‡After adjustment for multiple testing, p<0.02 is the threshold for statistical significance. §After adjustment for multiple testing, p<0.04 is the threshold for statistical significance. ||Ordinal outcome. ¶Continuous outcome.

### ***Table S5:* Symptom associations by severity of infection, among participants with test-confirmed previous SARS-CoV-2 infection**

|  | **Previous SARS-CoV-2 infection *vs* no infection** | | **SARS-CoV-2 infection >12 weeks prior *vs* no infection** | | **Previous SARS-CoV-2 infection *vs* previous non-COVID-19 ARI** | | **Previous non-COVID-19 ARI *vs*  no infection** | |
| --- | --- | --- | --- | --- | --- | --- | --- | --- |
|  | Estimate (95% CI)* | p value† | Estimate (95% CI)* | p value‡ | Estimate (95% CI)* | p value§ | Estimate (95% CI)* | p value\|\| |
| Coughing | 2.83 (2.06–3.89) | <0.001 | 1.57 (0.90–2.75) | 0.113 | 1.30 (0.85–1.97) | 0.225 | 2.92 (2.28–3.75) | <0.001 |
| Problems with sleep | 1.45 (1.13–1.86) | 0.003 | 1.64 (1.13–2.39) | 0.010 | 0.91 (0.64–1.31) | 0.625 | 1.48 (1.21–1.80) | <0.001 |
| Memory problems | 1.95 (1.44–2.65) | <0.001 | 1.75 (1.10–2.79) | 0.019 | 1.42 (0.93–2.17) | 0.100 | 1.65 (1.29–2.10) | <0.001 |
| Difficulty concentrating | 1.67 (1.23–2.27) | 0.001 | 1.42 (0.89–2.28) | 0.145 | 1.42 (0.95–2.12) | 0.090 | 1.51 (1.19–1.91) | <0.001 |
| Pains in muscles or joints | 1.38 (1.06–1.80) | 0.016 | 1.46 (0.98–2.17) | 0.060 | 1.09 (0.75–1.58) | 0.659 | 1.23 (1.00–1.51) | 0.055 |
| Problems with sense of smell / taste | 16.32 (11.62–22.94) | <0.001 | 13.94 (8.47–22.93) | <0.001 | 17.88 (9.06–35.29) | <0.001 | 1.33 (0.74–2.38) | 0.343 |
| Diarrhoea | 2.20 (1.49–3.24) | <0.001 | 1.48 (0.78–2.82) | 0.229 | 1.36 (0.82–2.26) | 0.240 | 2.08 (1.54–2.79) | <0.001 |
| Stomach (abdominal pains) | 1.65 (1.12–2.44) | 0.012 | 1.65 (0.94–2.91) | 0.082 | 0.87 (0.53–1.42) | 0.572 | 2.13 (1.63–2.79) | <0.001 |
| Changes to voice | 3.34 (2.04–5.46) | <0.001 | 2.63 (1.24–5.60) | 0.012 | 1.52 (0.82–2.83) | 0.184 | 3.16 (2.16–4.63) | <0.001 |
| Hair loss | 1.61 (1.02–2.55) | 0.040 | 1.44 (0.71–2.89) | 0.309 | 1.77 (0.89–3.49) | 0.101 | 0.94 (0.63–1.41) | 0.773 |
| Unusual racing of the heart | 2.38 (1.65–3.45) | <0.001 | 2.30 (1.33–3.97) | 0.003 | 1.34 (0.80–2.23) | 0.265 | 1.78 (1.32–2.40) | <0.001 |
| Lightheadedness or dizziness | 2.32 (1.68–3.20) | <0.001 | 2.27 (1.41–3.65) | <0.001 | 1.51 (0.96–2.39) | 0.075 | 1.54 (1.17–2.02) | 0.002 |
| Unusual sweating | 2.25 (1.51–3.34) | <0.001 | 2.20 (1.23–3.96) | 0.008 | 1.39 (0.79–2.42) | 0.252 | 1.82 (1.32–2.53) | <0.001 |
| MRC Dyspnoea¶ | 1.89 (1.45–2.47) | <0.001 | 1.55 (1.03–2.34) | 0.036 | 1.64 (1.14–2.37) | 0.008 | 1.36 (1.10–1.68) | 0.005 |
| PHQ-4 grade¶ | 1.34 (1.02–1.77) | 0.037 | 0.98 (0.63–1.53) | 0.941 | 0.98 (0.67–1.43) | 0.910 | 1.30 (1.05–1.60) | 0.017 |
| FACIT-13 score (reversed)** | 4.3% (2.4–6.1) | <0.001 | 2.1% (-0.3–4.6) | 0.092 | 1.8% (-1.3–4.9) | 0.263 | 3.4% (2.1–4.7) | <0.001 |
| EQ-5D Activities¶ | 2.60 (1.80–3.74) | <0.001 | 2.21 (1.27–3.83) | 0.005 | 1.36 (0.84–2.21) | 0.216 | 1.84 (1.38–2.44) | <0.001 |
| EQ-5D Pain¶ | 1.44 (1.09–1.91) | 0.009 | 1.58 (1.05–2.38) | 0.028 | 0.96 (0.65–1.41) | 0.823 | 1.40 (1.12–1.73) | 0.002 |
| EQ-5D Self-care (two groups) | 0.71 (0.29–1.71) | 0.440 | 0.95 (0.32–2.85) | 0.923 | 1.15 (0.41–3.26) | 0.788 | 0.93 (0.54–1.60) | 0.783 |
| EQ-5D Mobility (two groups) | 1.56 (1.03–2.37) | 0.035 | 1.64 (0.90–2.97) | 0.105 | 1.34 (0.77–2.34) | 0.307 | 1.19 (0.86–1.63) | 0.288 |
| EQ-5D VAS** | -2.7% (-4.7 to -0.7) | 0.007 | -1.9% (-4.8 to 1.0) | 0.199 | -0.6% (-3.6 to 2.4) | 0.700 | -1.7% (-3.2 to -0.1) | 0.036 |

Analysis is done in 228 participants with previous SARS-CoV-2 infection (202 symptomatic), 458 participants with previous non-COVID-19 ARI, and 8388 participants with no infection. ARI=acute respiratory infection. PHQ=Patient Health Questionnaire. MRC=Medical Research Council. VAS=visual analogue scale. *Estimates are odds ratios for binary and ordinal outcomes, and predicted percentage point changes for continuous outcomes. †After adjustment for multiple testing, p<0.048 is the threshold for statistical significance. ‡After adjustment for multiple testing, p<0.016 is the threshold for statistical significance. §After adjustment for multiple testing, p<0.002 is the threshold for statistical significance. ||After adjustment for multiple testing, p<0.04 is the threshold for statistical significance. ¶Ordinal outcome. **Continuous outcome.

### ***Table S6:* Symptom comparisons among participants with test-confirmed previous SARS-CoV-2 infection, non-COVID-19 ARIs, or no infection, adjusted for pre-infection general health**

|  | **SARS-CoV-2 infection >12 weeks prior *vs* non-COVID-19 ARI >12 weeks prior** | |
| --- | --- | --- |
|  | Estimate* (95% CI) | p value† |
| Coughing | 0.87 (0.57–1.31) | 0.498 |
| Problems with sleep | 0.79 (0.58–1.09) | 0.148 |
| Memory problems | 1.41 (0.96–2.08) | 0.079 |
| Difficulty concentrating | 1.33 (0.92–1.92) | 0.133 |
| Pains in muscles or joints | 1.16 (0.83–1.62) | 0.384 |
| Problems with sense of smell / taste | 7.91 (3.18–19.66) | <0.001 |
| Diarrhoea | 0.93 (0.58–1.51) | 0.779 |
| Stomach (abdominal pains) | 1.00 (0.64–1.54) | 0.984 |
| Changes to voice | 1.21 (0.64–2.30) | 0.551 |
| Hair loss | 2.22 (1.22–4.03) | 0.009 |
| Unusual racing of the heart | 1.32 (0.83–2.11) | 0.240 |
| Lightheadedness or dizziness | 1.36 (0.90–2.06) | 0.144 |
| Unusual sweating | 1.06 (0.66–1.68) | 0.818 |
| MRC Dyspnoea‡ | 1.06 (0.76–1.48) | 0.735 |
| PHQ-4 grade‡ | 0.92 (0.66–1.28) | 0.624 |
| FACIT-13 score (reversed)§ | 0.0% (-2.6 to 2.6) | 0.999 |
| EQ-5D Activities‡ | 1.20 (0.78–1.85) | 0.417 |
| EQ-5D Pain‡ | 1.02 (0.73–1.43) | 0.896 |
| EQ-5D Self-care (two groups) | 1.56 (0.67–3.61) | 0.300 |
| EQ-5D Mobility (two groups) | 0.88 (0.55–1.41) | 0.585 |
| EQ-5D VAS§ | 0.0% (-2.6 to 2.6) | 0.999 |

Analysis is done in 1131 participants with previous symptomatic SARS-CoV-2 infection and 198 participants with previous non-COVID-19 ARI. ARI=acute respiratory infection. PHQ=Patient Health Questionnaire. MRC=Medical Research Council. VAS=visual analogue scale. *Estimates are odds ratios for binary and ordinal outcomes, and predicted percentage point changes for continuous outcomes. †After adjustment for multiple testing, p<0.002 is the threshold for statistical significance. ‡Ordinal outcome. §Continuous outcome.

### ***Table S7:* Symptom associations by infection status, among participants with previous infection more than 12 weeks prior**

### **Latent class analysis**

In order to choose the optimal latent class model, we first fit models including only the chosen indicators with up to ten classes. The Vuong-Lo-Mendell-Rubin test showed significant improvement in model fit with almost all additional classes, but the BIC plot showed a clear inflection point at three classes, suggesting that addition of further classes did not lead to the same improvement in model fit.

| **Classes** | **BIC** | **AIC(LL)** | **AIC3(LL)** | **VLMR** | **p value** | **-2LL Diff** | **Bootstrapped p value** | **Classification errors** | **Entropy** |
| --- | --- | --- | --- | --- | --- | --- | --- | --- | --- |
| 1 | 17513.6 | 17388.7 | 17412.7 | ·· | ·· |  |  | 0.000 | 1.000 |
| 2 | 12402.5 | 12163.1 | 12209.1 | 5269.6 | <0.001 | 5269.6 | <0.001 | 0.023 | 0.908 |
| 3 | 8554.4 | 8200.7 | 8268.7 | 4006.5 | <0.001 | 4006.5 | <0.001 | 0.023 | 0.948 |
| 4 | 8057.2 | 7588.9 | 7678.9 | 655.7 | <0.001 | 655.7 | <0.001 | 0.047 | 0.911 |
| 5 | 7501.3 | 6918.6 | 7030.6 | 714.3 | <0.001 | 714.3 | <0.001 | 0.045 | 0.925 |
| 6 | 7323.1 | 6625.9 | 6759.9 | 336.7 | <0.001 | 336.7 | <0.001 | 0.055 | 0.912 |
| 7 | 6897.7 | 6086.1 | 6242.1 | 583.8 | <0.001 | 583.8 | <0.001 | 0.055 | 0.920 |
| 8 | 6816.7 | 5890.6 | 6068.6 | 239.5 | <0.001 | 239.5 | <0.001 | 0.058 | 0.917 |
| 9 | 6528.7 | 5488.2 | 5688.2 | 446.4 | <0.001 | 446.4 | <0.001 | 0.062 | 0.918 |
| 10 | 6701.9 | 5546.9 | 5768.9 | -14.8 | 0.962 | -14.8 | 1.000 | 0.060 | 0.917 |

BIC=Bayesian Information Criterion. AIC=Akaike Information Criterion. LL=log likelihood. VLMR= Vuong-Lo-Mendell-Rubin.

#### ***Table S8:* Fit statistics for initial latent class model for participants with previous SARS-CoV-2 infections**

SARS-CoV-2 infection model

BIC

| Number of classes | BIC |
| --- | --- |
| 1 | 17513.6 |
| 2 | 12402.5 |
| 3 | 8554.43 |
| 4 | 8057.16 |
| 5 | 7501.3 |
| 6 | 7323.06 |
| 7 | 6897.73 |
| 8 | 6816.66 |
| 9 | 6528.7 |
| 10 | 6701.94 |

BIC=Bayesian Information Criterion.

#### ***Figure S5:* BIC plot for initial latent class models**

Additionally, examination of the profile plots showed that the class separation was less distinct in models with more than three classes, causing problems with interpretation. We therefore chose a three-class model.

The three-class model had large bivariate residuals, showing that the latent classes were not fully explaining the covariance between the included indicators. We took two approaches to improve model fit: first, we examined all pairwise correlations between variables and sequentially included direct effects for all pairs of variables with an absolute correlation of 0.5 or more, starting with the variable pair with the highest correlation; second, we sequentially included direct effects according to largest bivariate residual between variables, retaining the direct effect in the model if it improved model fit. While model fit was significantly improved by these approaches, neither approach led to a complete reduction of all bivariate residuals. To ensure parsimony, we therefore examined the BIC for signs of an inflection point and, based on this assessment, retained one direct effect between memory problems and concentration.

SARS-CoV-2 infection model

BIC

| Number of direct effects | BIC(LL) |
| --- | --- |
| 0 | 8554.43 |
| 1 | 8357.78 |
| 2 | 8270.11 |
| 3 | 8178.32 |
| 4 | 8144.46 |
| 5 | 8111.22 |
| 6 | 8087.7 |
| 7 | 8056.18 |
| 8 | 8041.14 |
| 9 | 8022.55 |
| 10 | 8004.58 |

BIC=Bayesian Information Criterion.

#### ***Figure S6:* BIC plot for three-class SARS-CoV-2 models with direct effects**

Finally, to adjust for age and sex, we included each of these covariates individually in the model and examined their bivariate residuals with the indicator variables. We then added a direct effect for any pairs with a bivariate residual greater than 3.

The final model had high entropy (0.949) and showed well separated average latent class posterior probabilities.

We repeated the same procedure to establish latent class analysis models for previous non-COVID-19 ARIs and participants with no recorded infections. This led to a three-class model with two direct effects (between memory problems and concentration, and between stomach problems and diarrhoea) for non-COVID-19 ARIs, and a two-class model with one direct effect (between memory problems and concentration) for the no infection model.

|  | **SARS-CoV-2 model** | | | **Non-COVID-19 ARI model** | | | **No infection model** | |
| --- | --- | --- | --- | --- | --- | --- | --- | --- |
|  | Cluster1 | Cluster2 | Cluster3 | Cluster1 | Cluster2 | Cluster3 | Cluster1 | Cluster2 |
| Cluster1 | 1.000 | 0.000 | 0.000 | 0.999 | 0.000 | 0.000 | 1.000 | 0.000 |
| Cluster2 | 0.000 | 0.999 | 0.000 | 0.000 | 1.000 | 0.000 | 0.000 | 1.000 |
| Cluster3 | 0.000 | 0.000 | 1.000 | 0.001 | 0.000 | 1.000 | ·· | ·· |

#### ***Table S9:* Average latent class posterior probabilities of the final models**

Non-COVID-19 ARIs

BIC

| Number of classes | BIC(LL) |
| --- | --- |
| 1 | 6186.08 |
| 2 | 4917 |
| 3 | 4054.83 |
| 4 | 4008.28 |
| 5 | 3995.77 |
| 6 | 4015.58 |
| 7 | 3975.95 |
| 8 | 4046.35 |
| 9 | 4014.65 |
| 10 | 4159.99 |

No infections

| Number of classes | BIC(LL) |
| --- | --- |
| 1 | 56661.5 |
| 2 | 6241.01 |
| 3 | -5009 |
| 4 | -10889.3 |
| 5 | -14933.8 |
| 6 | -19409.7 |
| 7 | -25113 |
| 8 | -28382.2 |
| 9 | -30235 |
| 10 | -30925.3 |

BIC

| Number of direct effects | BIC(LL) |
| --- | --- |
| 0 | 3981.96 |
| 1 | 3956.41 |
| 2 | 3936.71 |
| 3 | 3927.77 |
| 4 | 3922.63 |
| 5 | 3912.58 |
| 6 | 3910.29 |
| 7 | 3906.19 |
| 8 | 3904.67 |
| 9 | 3902.86 |

| Number of direct effects | BIC(LL) |
| --- | --- |
| 0 | 6241.01 |
| 1 | 5073.98 |
| 2 | 4679.89 |
| 3 | 4246.66 |
| 4 | 3399.52 |
| 5 | 3129.19 |
| 6 | 2738.44 |
| 7 | 2593.07 |
| 8 | 2421.12 |
| 9 | 2079.61 |
| 10 | 1614.61 |

BIC=Bayesian Information Criterion.

#### ***Figure S7:* BIC plots for non-COVID-19 ARI and no infection models**

|  | **Mild  (45%)** | **Moderate (32%)** | **Severe (23%)** |
| --- | --- | --- | --- |
| Diarrhoea | 0.05 | 0.07 | 0.27 |
| Stomach problems | 0.05 | 0.10 | 0.34 |
| Pains in muscles or joints | 0.11 | 0.49 | 0.79 |
| FACIT-13† | 0.07 | 0.13 | 0.44 |
| Sleep problems | 0.25 | 0.44 | 0.76 |
| Memory problems | 0.08 | 0.19 | 0.69 |
| Difficulty concentrating | 0.07 | 0.19 | 0.76 |
| Unusual racing of the heart | 0.04 | 0.09 | 0.42 |
| Lightheadedness or dizziness | 0.06 | 0.15 | 0.54 |
| Unusual sweating | 0.04 | 0.09 | 0.36 |
| MRC Dyspnoea‡ | 0.09 | 0.18 | 0.44 |
| Coughing | 0.09 | 0.14 | 0.33 |
| Changes to voice | 0.03 | 0.03 | 0.20 |
| Hair loss | 0.06 | 0.09 | 0.25 |
| Problems with smell/taste | 0.09 | 0.11 | 0.40 |
| EQ-5D VAS† | 0.19 | 0.27 | 0.44 |
| EQ-5D Utility Index† | 0.00 | 0.13 | 0.24 |
| PHQ-4 grade‡ | 0.02 | 0.13 | 0.37 |

MRC =Medical Research Council. VAS=visual analogue scale. PHQ=Patient Health Questionnaire. *Conditional probability is shown for binary variables, and mean score for ordinal and continuous variables. †Continuous variables have been reversed to aid with interpretation, so that higher values indicate worse severity or health state. ‡Ordinal variable.

### ***Table S10:* Conditional probabilities or mean severity scores for all symptoms among participants with previous SARS-CoV-2 infection**

|  | **Mild (N=608)** | **Moderate (N=432)** | **Severe (N=303)** |
| --- | --- | --- | --- |
| **Sociodemographics** | | | |
| Age, years | 59.3 (50.8-65.9) | 61.4 (53.5-67.5) | 56.2 (49.0-64.3) |
| <30 | 24 (3.9%) | 13 (3.0%) | 6 (2.0%) |
| 30 to <40 | 40 (6.6%) | 26 (6.0%) | 20 (6.6%) |
| 40 to <50 | 78 (12.8%) | 41 (9.5%) | 62 (20.5%) |
| 50 to <60 | 179 (29.4%) | 116 (26.9%) | 97 (32.0%) |
| 60 to <70 | 207 (34.0%) | 172 (39.8%) | 88 (29.0%) |
| ≥70 | 80 (13.2%) | 64 (14.8%) | 30 (9.9%) |
| Sex |  |  |  |
| Female | 411 (67.6%) | 294 (68.1%) | 219 (72.3%) |
| Male | 197 (32.4%) | 138 (31.9%) | 84 (27.7%) |
| Ethnicity |  |  |  |
| White | 575 (94.6%) | 405 (93.8%) | 279 (92.1%) |
| Mixed/multiple/other ethnic groups | 19 (3.1%) | 16 (3.7%) | 11 (3.6%) |
| South Asian | 10 (1.6%) | 6 (1.4%) | 11 (3.6%) |
| Black/African/Caribbean/Black British | 4 (0.7%) | 5 (1.2%) | 2 (0.7%) |
| Country |  |  |  |
| England | 557 (91.6%) | 376 (87.0%) | 274 (90.4%) |
| Northern Ireland | 7 (1.2%) | 7 (1.6%) | 6 (2.0%) |
| Scotland | 31 (5.1%) | 30 (6.9%) | 12 (4.0%) |
| Wales | 13 (2.1%) | 19 (4.4%) | 11 (3.6%) |
| No. people per bedroom |  |  |  |
| <1 | 389 (64.4%) | 288 (66.8%) | 149 (49.8%) |
| 1 to <2 | 203 (33.6%) | 132 (30.6%) | 137 (45.8%) |
| 2 to <3 | 11 (1.8%) | 11 (2.6%) | 11 (3.7%) |
| 3 to <4 | 1 (0.2%) | 0 | 2 (0.7%) |
| Quartiles of IMD decile |  |  |  |
| Q4 (least deprived) | 214 (35.3%) | 136 (31.5%) | 81 (26.7%) |
| Q3 | 132 (21.7%) | 104 (24.1%) | 63 (20.8%) |
| Q2 | 129 (21.3%) | 87 (20.1%) | 60 (19.8%) |
| Q1 (most deprived) | 132 (21.7%) | 105 (24.3%) | 99 (32.7%) |
| Frontline worker |  |  |  |
| No | 526 (86.5%) | 350 (81.0%) | 215 (71.0%) |
| Non-health | 61 (10.0%) | 67 (15.5%) | 62 (20.5%) |
| Health | 21 (3.5%) | 15 (3.5%) | 26 (8.6%) |
| Highest educational level attained |  |  |  |
| Primary or secondary | 51 (8.4%) | 60 (13.9%) | 47 (15.5%) |
| Higher or further (A levels) | 84 (13.8%) | 62 (14.4%) | 41 (13.5%) |
| College or university | 282 (46.4%) | 191 (44.2%) | 132 (43.6%) |
| Post-graduate | 191 (31.4%) | 119 (27.5%) | 83 (27.4%) |
| **Clinical characteristics** | | | |
| BMI, kg/m^2^ | 24.7 (22.3-27.5) | 26.0 (23.5-29.8) | 27.4 (24.0-31.3) |
| <25 | 325 (53.5%) | 181 (41.9%) | 97 (32.0%) |
| 25 to <30 | 203 (33.4%) | 148 (34.3%) | 104 (34.3%) |
| ≥30 | 79 (13.0%) | 103 (23.8%) | 102 (33.7%) |
| Asthma | 91 (15.0%) | 95 (22.0%) | 83 (27.4%) |
| Atopy | 156 (25.7%) | 116 (26.9%) | 87 (28.7%) |
| Autoimmune disease | 36 (5.9%) | 40 (9.3%) | 37 (12.2%) |
| Cancer |  |  |  |
| Past (cured or in remission) | 51 (8.4%) | 39 (9.0%) | 17 (5.6%) |
| Active treatment | 3 (0.5%) | 6 (1.4%) | 1 (0.3%) |
| COPD | 12 (2.0%) | 12 (2.8%) | 14 (4.6%) |
| Diabetes | 5 (0.8%) | 15 (3.5%) | 22 (7.3%) |
| Heart disease | 7 (1.2%) | 16 (3.7%) | 13 (4.3%) |
| Hypertension | 93 (15.3%) | 98 (22.7%) | 69 (22.8%) |
| Immunodeficiency | 6 (1.0%) | 2 (0.5%) | 5 (1.7%) |
| Kidney disease | 11 (1.8%) | 4 (0.9%) | 11 (3.6%) |
| Major neurological conditions | 16 (2.6%) | 13 (3.0%) | 11 (3.6%) |
| Number of symptoms reported* | 1 (0-2) | 3 (1-4) | 8 (6-10) |
| **Infections** | | | |
| Weeks since infection |  |  |  |
| 4–12 weeks | 80 (13.2%) | 42 (9.7%) | 44 (14.5%) |
| >12 weeks | 528 (86.8%) | 390 (90.3%) | 259 (85.5%) |
| Infection severity† |  |  |  |
| Asymptomatic | 46/318 (14.5%) | 38/235 (16.2%) | 6/204 (2.9%) |
| Mildly unwell | 85/318 (26.7%) | 44/235 (18.7%) | 32/204 (15.7%) |
| Moderately unwell | 82/318 (25.8%) | 53/235 (22.6%) | 43/204 (21.1%) |
| Very unwell | 99/318 (31.1%) | 94/235 (40.0%) | 95/204 (46.6%) |
| Hospitalised | 6/318 (1.9%) | 6/235 (2.6%) | 28/204 (13.7%) |
| Ever reported long COVID‡ | 81 (13.3%) | 93 (21.5%) | 168 (55.4%) |
| Reported long COVID in current questionnaire | 38 (6.3%) | 48 (11.1%) | 144 (47.5%) |

Data are n (%) or median (IQR). BMI=body-mass index. IMD=Index of Multiple Deprivation. *Does not include health-related quality of life measurements. †For symptomatic and non-hospitalised participants, severity was self-reported with the following statements: “Mildly unwell – I could do most of my usual activities”, “Moderately unwell – I couldn’t do usual activities but didn’t need to go to bed in the daytime”, and “Very unwell – I had to go to bed in the daytime”. ‡Answered ‘Yes’ to the question “Would YOU say that you currently have 'long COVID', i.e. ongoing symptoms more than four weeks after the onset of proven or suspected SARS-CoV-2 infection” before or on date of survey.

### ***Table S11:* Participant characteristics by symptom cluster for SARS-CoV-2 model**

|  | **Previous SARS-CoV-2 infection (N=303)** | **Previous non-COVID-19 ARI (N=101)** | **No infection (N=3,664)** |
| --- | --- | --- | --- |
| **Sociodemographics** | | | |
| Age, years | 56.2 (49.0-64.3) | 55.2 (42.0-63.5) | 63.6 (54.7-69.5) |
| <30 | 6 (2.0%) | 12 (11.9%) | 119 (3.2%) |
| 30 to <40 | 20 (6.6%) | 11 (10.9%) | 195 (5.3%) |
| 40 to <50 | 62 (20.5%) | 18 (17.8%) | 334 (9.1%) |
| 50 to <60 | 97 (32.0%) | 25 (24.8%) | 724 (19.8%) |
| 60 to <70 | 88 (29.0%) | 25 (24.8%) | 1,450 (39.6%) |
| ≥70 | 30 (9.9%) | 10 (9.9%) | 842 (23.0%) |
| Sex |  |  |  |
| Female | 219 (72.3%) | 81 (80.2%) | 2,628 (71.7%) |
| Male | 84 (27.7%) | 20 (19.8%) | 1,036 (28.3%) |
| Ethnicity |  |  |  |
| White | 279 (92.1%) | 94 (93.1%) | 3,505 (95.7%) |
| Mixed/multiple/other ethnic groups | 11 (3.6%) | 4 (4.0%) | 87 (2.4%) |
| South Asian | 11 (3.6%) | 2 (2.0%) | 49 (1.3%) |
| Black/African/Caribbean/Black British | 2 (0.7%) | 1 (1.0%) | 23 (0.6%) |
| Country |  |  |  |
| England | 274 (90.4%) | 90 (89.1%) | 3,216 (87.8%) |
| Northern Ireland | 6 (2.0%) | 4 (4.0%) | 63 (1.7%) |
| Scotland | 12 (4.0%) | 5 (5.0%) | 231 (6.3%) |
| Wales | 11 (3.6%) | 2 (2.0%) | 153 (4.2%) |
| No. people per bedroom |  |  |  |
| <1 | 149 (49.8%) | 47 (47.0%) | 2,480 (68.1%) |
| 1 to <2 | 137 (45.8%) | 49 (49.0%) | 1,090 (29.9%) |
| 2 to <3 | 11 (3.7%) | 3 (3.0%) | 68 (1.9%) |
| 3 to <4 | 2 (0.7%) | 1 (1.0%) | 2 (0.1%) |
| Quartiles of IMD decile |  |  |  |
| Q4 (least deprived) | 81 (26.7%) | 32 (31.7%) | 1,088 (29.7%) |
| Q3 | 63 (20.8%) | 24 (23.8%) | 969 (26.5%) |
| Q2 | 60 (19.8%) | 23 (22.8%) | 769 (21.0%) |
| Q1 (most deprived) | 99 (32.7%) | 22 (21.8%) | 837 (22.9%) |
| Frontline worker |  |  |  |
| No | 215 (71.0%) | 80 (79.2%) | 3,189 (87.2%) |
| Non-health | 62 (20.5%) | 17 (16.8%) | 384 (10.5%) |
| Health | 26 (8.6%) | 4 (4.0%) | 84 (2.3%) |
| Highest educational level attained |  |  |  |
| Primary or secondary | 47 (15.5%) | 18 (17.8%) | 477 (13.0%) |
| Higher or further (A levels) | 41 (13.5%) | 13 (12.9%) | 594 (16.2%) |
| College or university | 132 (43.6%) | 42 (41.6%) | 1,592 (43.5%) |
| Post-graduate | 83 (27.4%) | 28 (27.7%) | 995 (27.2%) |
| **Clinical characteristics** | | | |
| BMI, kg/m^2^ | 27.4 (24.0-31.3) | 27.1 (23.4-29.9) | 25.7 (23.0-29.7) |
| <25 | 97 (32.0%) | 38 (37.6%) | 1,609 (44.0%) |
| 25 to <30 | 104 (34.3%) | 38 (37.6%) | 1,173 (32.1%) |
| ≥30 | 102 (33.7%) | 25 (24.8%) | 872 (23.9%) |
| Asthma | 83 (27.4%) | 27 (26.7%) | 629 (17.2%) |
| Atopy | 87 (28.7%) | 33 (32.7%) | 943 (25.7%) |
| Autoimmune disease | 37 (12.2%) | 15 (14.9%) | 426 (11.6%) |
| Cancer |  |  |  |
| Past (cured or in remission) | 17 (5.6%) | 6 (5.9%) | 332 (9.1%) |
| Active treatment | 1 (0.3%) | 1 (1.0%) | 39 (1.1%) |
| COPD | 14 (4.6%) | 3 (3.0%) | 101 (2.8%) |
| Diabetes | 22 (7.3%) | 9 (8.9%) | 224 (6.1%) |
| Heart disease | 13 (4.3%) | 33 (32.7%) | 154 (4.2%) |
| Hypertension | 69 (22.8%) | 15 (14.9%) | 935 (25.5%) |
| Immunodeficiency | 5 (1.7%) | 1 (1.0%) | 29 (0.8%) |
| Kidney disease | 11 (3.6%) | 6 (5.9%) | 87 (2.4%) |
| Major neurological conditions | 11 (3.6%) | 2 (2.0%) | 124 (3.4%) |
| Number of symptoms reported* | 8 (6-10) | 7 (5-9) | 2 (1-4) |
| **Infections** | | | |
| Weeks since infection |  |  |  |
| 4–12 weeks | 44 (14.5%) | 59 (58.4%) | ·· |
| >12 weeks | 259 (85.5%) | 42 (41.6%) | ·· |
| Infection severity† |  |  |  |
| Asymptomatic | 6/204 (2.9%) | ·· | ·· |
| Mildly unwell | 32/204 (15.7%) | 42 (41.6%) | ·· |
| Moderately unwell | 43/204 (21.1%) | 31 (30.7%) | ·· |
| Very unwell | 95/204 (46.6%) | 22 (21.8%) | ·· |
| Hospitalised | 28/204 (13.7%) | 6 (5.9%) | ·· |
| Ever reported long COVID‡ | 168 (55.4%) | 17 (16.8%) | 64 (1.75%) |
| Reported long COVID in current questionnaire§ | 144 (47.5%) | 12 (11.9%) | 41 (1.1%) |

Data are n (%) or median (IQR). ARI=acute respiratory infection. BMI=body-mass index. IMD=Index of Multiple Deprivation. * Does not include health-related quality of life measurements. †For symptomatic and non-hospitalised participants, severity was self-reported with the following statements: “Mildly unwell – I could do most of my usual activities”, “Moderately unwell – I couldn’t do usual activities but didn’t need to go to bed in the daytime”, and “Very unwell – I had to go to bed in the daytime”. ‡Answered ‘Yes’ to the question “Would YOU say that you currently have 'long COVID', i.e. ongoing symptoms more than four weeks after the onset of proven or suspected SARS-CoV-2 infection” before or on date of survey. §Previous non-COVID-19 ARI: 5 (2.6%) in the mild cluster and 3 (1.7%) in the moderate cluster reported long COVID. No infection: 14 (0.3%) in the mild cluster reported long COVID.

### ***Table S12:* Participant characteristics for the most severe symptom clusters, by infection status**
